# Novel mutation leading to splice donor loss in a conserved site of *DMD* causes cryptorchidism

**DOI:** 10.1101/2024.01.29.24301909

**Authors:** Jianhai Chen, Yangying Jia, Jie Zhong, Kun Zhang, Hongzheng Dai, Guanglin He, Fuping Li, Li Zeng, Chuanzhu Fan, Huayan Xu

## Abstract

**Background:** As one of the most common congenital abnormalities in male births, cryptorchidism has been found to have a polygenic etiology according to previous studies of common variants. However, little is known about genetic predisposition of rare variants for cryptorchidism, since rare variants have larger effective size on diseases than common variants.

**Methods:** In this study, a cohort of 115 Chinese probands with cryptorchidism was analyzed using whole-genome sequencing (WGS), alongside 19 parental controls and 2136 unaffected men. Additionally, CRISPR-Cas9 editing of a conserved variant was performed in a mouse model, with MRI screening utilized to observe the phenotype.

**Results:** In 30 of 115 patients (26.1%), we identified four novel genes (*ARSH*, *DMD*, *MAGEA4*, and *SHROOM2*) affecting at least five unrelated patients and four known genes (*USP9Y*, *UBA1*, *BCORL1*, and *KDM6A*) with the candidate rare pathogenic variants affecting at least two cases. Burden tests of rare variants revealed the genome-wide significances for newly identified genes (*p* < 2.5×10^-6^) under the Bonferroni correction. Surprisingly, novel and known genes were mainly from X chromosome (seven on X and one on Y) and all rare X-chromosomal segregating variants exhibited a maternal inheritance rather than *de novo* origin. CRISPR-Cas9 mouse modeling of a splice donor loss variant in *DMD* (NC_000023.11:g.32454661C>G), which resides in a conserved site across vertebrates, replicated bilateral cryptorchidism phenotypes, confirmed by Magnetic resonance imaging (MRI) at 4 and 10 weeks.

**Conclusion:** Our results revealed the role of the *DMD* gene mutation in causing cryptorchidism. The results also suggest that maternal-X inheritance of pathogenic defects could have a predominant role in the development of cryptorchidism.

## Introduction

Cryptorchidism (also known as ’hidden testicle’, OMIM 219050) has a global prevalence rate of 2% to 4% at full-term birth in boys ^1^. Despite its relatively low prevalence, cryptorchidism is the most common birth defect involving urogenital abnormalities in newborn boys. It is also the best-characterized risk factor for infertility and testicular cancer ^2^. Normally, the testes spontaneously descend into the scrotum by the eighth month (33 weeks) of pregnancy, likely triggered by transient activation of the hypothalamus-pituitary-gonadal (HPG) axis, leading to an increase in reproductive hormone levels ^3^ ^4^. In patients with cryptorchidism, one or both testes may fail to descend completely into the dependent part of the scrotum, resulting in unilateral or bilateral cryptorchidism, respectively.

Maternal risk factors before and during pregnancy, such as maternal smoking, endocrine-disrupting chemicals, and gestational diabetes, have increasingly been recognized as the risk factors for congenital cryptorchidism ^5^. However, the studies on genetic risk factors of cryptorchidism are still limited. Multiple etiological features, such as familial aggregation and increased prevalence in first-degree relatives, suggest a genetic predisposition to this disease ^6^. For instance, a family history of cryptorchidism can increase the risk of the condition in newborn boys by approximately threefold ^7^ ^8^. The recurrence risk has been found to be more than twice as high in brothers of cryptorchidism cases compared to healthy controls ^9^. Thus, despite cryptorchidism’s complex susceptibility to maternal and environmental factors, strong genetic components may also contribute to its etiology.

Previous microarray studies based on common variants (GWAS) have suggested a potential association of certain pathways with cryptorchidism but failed to identify specific genes, likely due to the small effect size of common variants ^10^. Indeed, both population genetics theory and empirical data suggest that rare variants have much larger effect sizes than common variants ^11–14^, indicating their significant role in causing genetic diseases. It is estimated that most human rare protein-altering variants (missense) are pathogenic ^15^. Furthermore, disruptive mutations (nonsense) are disproportionately common causal factors in about 11.5% of human genetic diseases ^16^. At the population level, these causal variants are rare in allele frequency due to strong purifying selection, which limits their accumulation across generations. Therefore, rare deleterious variants with large genetic effect sizes represent a promising area for discovering causative genes for genetic diseases ^17^, especially for diseases affecting fertility, such as cryptorchidism.

The application of various genetic screening techniques, including whole-genome and exome sequencing (WGS/WES), has facilitated the precise diagnosis of numerous genetic diseases caused by rare variants ^18–25^. For instance, a WES study on an Indian family identified a rare deleterious homozygous missense variant in the *RXFP2* gene, which causes cryptorchidism ^26^. Although WES is more commonly used than WGS due to its lower cost, WGS offers a more uniform distribution of sequencing quality (including single-nucleotide variants, insertions, and deletions) and a higher discovery rate of coding-region variants, approximately 3%^27^ ^28^.

To date, a rare variants study of cryptorchidism based on WGS is still lacking. In this study, we conducted rare variants screening based on WGS data for 134 Han Chinese samples including cases (115) and family controls (19). We also used a local cohort of 2136 unaffected men for comparison. We identified candidate rare pathogenic variants based on autosomal recessive, compound heterozygous, and X-linked recessive inheritance mode for cryptorchidism patients. We annotated these variants and recognized both known and novel candidate genes and variants. We also confirmed the role of a noval rare mutation in *DMD* (NC_000023.11:g.32454661C>G) in causing cryptorchidism based on transgenic mouse modeling. Our study may facilitate the development of molecular diagnosis and precision medicine for patients suffering from cryptorchidism.

## Results

### The MRI on typical bilateral and unilateral cryptorchidism and basic sequencing statistics

The diagnosis of cryptorchidism was based on a physical exam by professional pediatric urologists following standard protocols ^29^. Typical bilateral cryptorchidism and unilateral cryptorchidism of two patients were shown using the magnetic resonance imaging (MRI) (Figure 1a-1d). In comparison to controls, the patient with bilateral cryptorchidism had two non-descended testes, while the patient with unilateral cryptorchidism had only one undescended testis (Figure 1).

**Figure 1.**
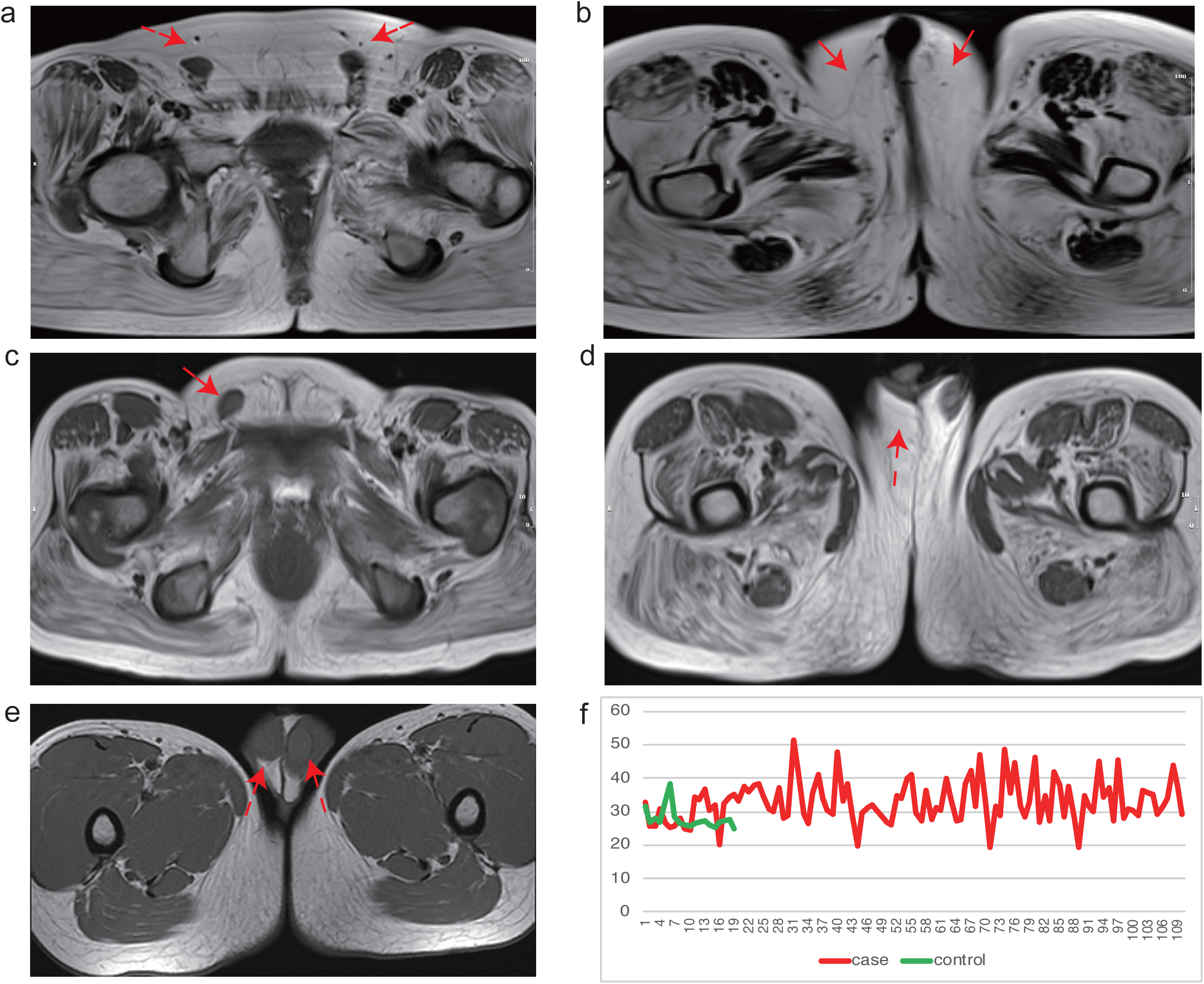
The Magnetic Resonance Imaging (MRI) and Sequencing Depth. (a-e) The two magnetic resonance imaging (MRI) graphs for bilateral cryptorchidism (case B130). (c-d) The two MRI graphs for unilateral cryptorchidism (case U111). (e) The MRI for one control. The red arrows show the location of the testes. (f) The WGS overall depths for the cases (red) and control (green).

We conducted WGS on 115 cases and 19 family controls. Among the cases, there were 21 (18.26%) bilateral cryptorchidism patients and 94 (81.74%) unilateral cryptorchidism patients. This dataset also includes three family trios of bilateral cryptorchidism, four trios of unilateral cryptorchidism, and three duos with only cryptorchidism cases and mothers. Based on previous sensitivity tests, 15x WGS depth/fold can achieve accurate SNV calling, while 30x is sufficient for calling indels ^30^ ^31^. In this study, the average sequencing depths were 33.03-fold for cases and 27.97-fold for controls, suggesting a balanced design of sequencing depth and quality for all samples (Supplementary Table 1, Figure 1f and Supplementary Figure 1a). To confirm the reported relationship, we used whole-genome SNPs to estimate the relatedness between family members. We also confirmed the population ancestry of all samples based on the principal component analysis (PCA). Based on identity by state (IBS) distances among family members, we confirmed that family members are genetically closer than unrelated individuals (Supplementary Figure 1b). We also confirmed that all parents-son relationships are within the coefficient range of first-degree relatives (from 0.177 to 0.354). By incorporating individuals from the “1000 genomes” project, we further revealed that all newly sequenced samples are closely related to the East Asian population (Supplementary Figure 1c).

### The Rare Candidate Pathogenic Variants for Known Genes

With the keywords ‘cryptorchidism variants’ and ‘cryptorchidism genetics’, we conducted the initial screening for known genes in the PubMed literature database ^32^. We retrieved 56 genes previously reported to be associated with cryptorchidism (Supplementary Table 2). Most of these previous studies were based on the genetic screening of small patient samples. Thus, it is interesting to know whether these genes can be confirmed in our cohort. We found that 41 of these genes were also registered in the Online Mendelian Inheritance in Man (OMIM) and Human Phenotype Ontology (HPO) database (June-2022) as genes related to cryptorchidism. We also added 738 genes associated with cryptorchidism phenotypes from these databases. These 779 genes (738+41) were used as a “pool” of known genes (Supplementary Table 2). We annotated non-redundant variants using the online tool VEP in the Ensembl database ^33^ and then summarized both known and novel candidate genes.

We detected four known genes (*BCORL1, KDM6A, UBA1,* and *USP9Y*) with candidate rare pathogenic variants present in at least two cases but absent in paternal controls (Table 1 and Supplementary Table 3). Six variants were not detected in all publicly available population databases (gnomAD, 1000 genomes, dbSNP, ExAC, ClinVar, etc.). One variant, *BCORL1* (NC_000023.11:g.130028727G>A;NP_001171701.1:p.(G1391R)), was also classified as rare based on the allele frequency of all human populations, with the highest frequency in East Asian population (0.005439; Supplementary Table 3). Although this allele frequency is relatively higher in the East Asian population than globally, it remains much lower than the global prevalence rate of cryptorchidism in boys, which is 2%-4%.

**Table 1.** The candidate pathogenic variants of known genes. “B” and “U” in ID column are “bilateral cryptorchidism” and “unilateral cryptorchidism”, respectively. The “Pred” column shows the number of prediction methods that support deleterious effects of the variants (see Supplementary Table 3 for specific methods). Note: *<u>USP9Y</u>* (underlined) is a Y chromosome gene, while all other genes reside on the X chromosome.

| ID | HVGS | Gene,Chr | Pred |
| --- | --- | --- | --- |
| U106 | NC_000023.11:g.130028727G>A;NP_001171701.1:p.(G1391R) | *BCORL1* | 6 |
| U37 | NC_000023.11:g.130028727G>A;NP_001171701.1:p.(G1391R) | *BCORL1* | 6 |
| U50 | NC_000023.11:g.130028727G>A;NP_001171701.1:p.(G1391R) | *BCORL1* | 6 |
| U9 | NC_000023.11:g.130028727G>A;NP_001171701.1:p.(G1391R) | *BCORL1* | 6 |
| U60 | NC_000023.11:g.45069958A>C;NP_001278345.1:p.(K775T) | *KDM6A* | 5 |
| U58 | NC_000023.11:g.45110152A>G;NP_001278344.1:p.(D1412G) | *KDM6A* | 6 |
| U51 | NC_000023.11:g.47212835A>T;NP_003325.2:p.(Y873F) | *UBA1* | 11 |
| U60 | NC_000023.11:g.47214843G>C;NP_695012.1:p.(V1031L) | *UBA1* | 5 |
| U49 | NC_000024.10:g.12722148G>A;NP_004645.2:p.(E96K) | <u>USP9Y</u> | 6 |
| U50 | NC_000024.10:g.12722148G>A;NP_004645.2:p.(E96K) | <u>USP9Y</u> | 7 |
| U109 | NC_000024.10:g.12739592C>G;NP_004645.2:p.(S462C) | <u>USP9Y</u> | 5 |
| U73 | NC_000024.10:g.12739592C>G;NP_004645.2:p.(S462C) | <u>USP9Y</u> | 5 |
| U13 | NC_000024.10:g.12739592C>G;NP_004645.2:p.(S462C) | <u>USP9Y</u> | 5 |
| U71 | NC_000024.10:g.12739592C>G;NP_004645.2:p.(S462C) | <u>USP9Y</u> | 5 |

Interestingly, all known genes affecting at least two cases are on sex chromosomes. Not a single autosomal gene was associated with more than one case. This strongly suggests a predominant role of sex chromosomal genes in cryptorchidism pathogenicity. The Y-chromosome gene *USP9Y* (Ubiquitin Specific Peptidase 9 Y-Linked) was detected in six boys with unilateral cryptorchidism. In addition, three X-chromosome genes (*BCORL1*, *UBA1*, and *KDM6A*) were found to carry candidate rare pathogenic variants affecting at least two patients. Strikingly, the two rare pathogenic variants in *BCORL1* (NC_000023.11:g.130028727G>A; NP_001171701.1:p.(G1391R)) and *USP9Y* (NC_000024.10:g.12739592C>G; NP_004645.2:p.(S462C)) were found in four unilateral patients per variant, with only the former registered in the gnomAD database. Both variants were predicted to be deleterious by multiple *in silico* algorithms (Supplementary Table 3). In addition, the wild-type alleles of these variants showed evolutionary conservation in primate species, based on sequence alignments of primate orthologous genes from the Ensembl database (v105, Supplementary Figure 2). Together, the candidate rare pathogenic variants of known genes were found to affect 12 cases, indicating a molecular diagnostic rate of 10.4% (12/115). The diagnostic rates for bilateral and unilateral cryptorchidism were 0% (0/21) and 12.77% (12/94), respectively. These candidate pathogenic variants and genotypes in cases and parents were validated using the Sanger Sequencing (Supplementary Figure 3).

### The Rare Candidate Pathogenic Variants for Novel Genes

To identify candidate pathogenic variants in novel candidate genes for cryptorchidism, we excluded cases with the above-mentioned pathogenic variants in known genes. Subsequently, we ranked genes based on the burden of rare deleterious variants that occurred in cases. To increase the reliability of the newly identified genes, we focused on genes affecting at least five cases with candidate variants. We found four genes that fulfilled these rigorous requirements: *DMD*, *ARSH*, *MAGEA4*, and *SHROOM2* (Table 2).

**Table 2.** The pathogenic variants in novel candidate genes. “B” and “U” in the ID column are “bilateral cryptorchidism” and “unilateral cryptorchidism”, respectively. The “Pred” column shows the number of prediction methods supporting the variants as deleterious (see Supplementary Table 4 for specific *in silico* prediction algorithms). Note: all genes are on the X chromosome.

| ID | HGVSc | Gene | Pred |
| --- | --- | --- | --- |
| U27 | NC_000023.11:g.3024049G>C;NP_001011719.1:p.(E310D) | ARSH | 5 |
| U64 | NC_000023.11:g.3024093G>C;NP_001011719.1:p.(G325A) | ARSH | 13 |
| B122 | NC_000023.11:g.3033050C>G;NP_001011719.1:p.(P452A) | ARSH | 9 |
| U127 | NC_000023.11:g.3033050C>G;NP_001011719.1:p.(P452A) | ARSH | 9 |
| U28 | NC_000023.11:g.3033050C>G;NP_001011719.1:p.(P452A) | ARSH | 9 |
| U42 | NC_000023.11:g.31774145C>T; LRG_199p1:p.(E2453K) | DMD | 3 |
| U55 | NC_000023.11:g.32342234G>A;NP_004000.1:p.(R1926C) | DMD | 6 |
| U114 | NC_000023.11:g.32346044G>C;NP_000100.2:p.(Q1821E) | DMD | 1 |
| U58 | NC_000023.11:g.32346044G>C;NP_000100.2:p.(Q1821E) | DMD | 1 |
| B130 | NC_000023.11:g.32454661C>G (splice donor loss variant) | DMD | 1 |
| U70 | NC_000023.11:g.32472199A>G;NP_000100.2:p.(Y964H) | DMD | 5 |
| U111 | NC_000023.11:g.32573783C>T; NP_000100.2:p.(D548N) | DMD | 7 |
| U106 | NC_000023.11:g.151924064G>A;NP_001011550.1:p.(A134T) | MAGEA4 | 2 |
| U36 | NC_000023.11:g.151924064G>A;NP_001011550.1:p.(A134T) | MAGEA4 | 2 |
| U8 | NC_000023.11:g.151924064G>A;NP_001011550.1:p.(A134T) | MAGEA4 | 2 |
| U94 | NC_000023.11:g.151924064G>A;NP_001011550.1:p.(A134T) | MAGEA4 | 2 |
| U27 | NC_000023.11:g.151924194A>T;NP_001373127.1:p.(Y177F) | MAGEA4 | 4 |
| U98 | NC_000023.11:g.9786618C>A;NP_001640.1:p.(R25S) | SHROOM2 | 2 |
| U33 | NC_000023.11:g.9894590G>A;NP_001640.1:p.(D228N) | SHROOM2 | 2 |
| U2 | NC_000023.11:g.9937394C>T;NP_001307593.1:p.(P118L) | SHROOM2 | 5 |
| U55 | NC_000023.11:g.9937537G>T;NP_001307593.1:p.(A166S) | SHROOM2 | 2 |
| B79 | NC_000023.11:g.9937537G>T;NP_001307593.1:p.(A166S) | SHROOM2 | 5 |
| B81 | NC_000023.11:g.3024049G>C;NP_001011719.1:p.(E310D) | SHROOM2 | 5 |

Based on allele frequencies in multiple genome databases (gnomAD, 1000 genomes, dbSNP, ExAC, ClinVar, etc.) and *in silico* predictions, all variants were classified as ‘rare’ based on allele frequencies in normal populations (allele frequency < 0.01) and classified as ‘deleterious’ by at least one prediction method. The candidate pathogenic variants of these novel genes were found in 20 cases but not in paternal controls, nor in a local cohort of 2136 healthy men (Supplementary Table 4). Moreover, the wild-type alleles showed a primate-wide evolutionary conservation based on the Ensembl sequence alignments of orthologous genes (Supplementary Figure 4). Interestingly, a splice donor variant in *DMD* (NC_000023.11:g.32454661C>G) is located in a site highly conserved across vertebrates, ranging from mammals to birds, reptiles, and fish (Figure 3). The cases U27 and U55 both had two candidate variants. Among the affected cases, four were bilateral and involved in genes *ARSH*, *DMD*, and *SHROOM2*. The remaining 16 unilateral cases were affected by all four genes. Thus, the diagnostic rates for bilateral and unilateral cryptorchidism are 19.05% (4/21) and 17.02% (16/94), respectively. Together with the known genes, the overall diagnostic rates are 23.81% (5/21) and 28.72% (27/94) for bilateral and unilateral cryptorchidism, respectively.

Based on gnomAD records, five variants showed low allele frequencies in both the global and East Asian population, while the remaining ten variants were newly discovered without information on allele frequency in databases (Supplementary Table 4). We also found recurrent variants in multiple cases: NC_000023.11:g.32346044G>C; NP_000100.2:p.(Q1821E) (*DMD*, two patients), NC_000023.11:g.3033050C>G; NP_001011719.1:p.(P452A) (*ARSH*, three patients), NC_000023.11:g.151924064G>A; NP_001011550.1:p.(A134T) (*MAGEA4*, four patients), and NC_000023.11:g.9937537G>T; NP_001307593.1:p.(A166S) (*SHROOM2*, two patients). These candidate variants and genotypes were also validated using the Sanger Sequencing for all samples (Supplementary Figure 5).

### Burden Tests Support the Statistical Significance of Novel Candidate genes

To statistically test our newly identified candidate genes, we performed burden tests for rare variants with the RVTEST package ^34^. We further used Bonferroni correction to account for multiple testing (p = 2.5×10^-6^). We found that five tests supported the whole-genome significance of three genes *DMD*, *MAGEA4*, and *SHROOM2* (Table 3, p < 2.5×10^-6^). For *ARSH*, four methods, except for the CMC Wald test, supported the genome-wide significance after Bonferroni correction (p < 2.5×10^-6^).

**Table 3.** The significance levels of burden tests for novel candidate genes with rare variants.

| Gene | CMC | CMCFisherExact | CMCWald | Fp | Zeggini |
| --- | --- | --- | --- | --- | --- |
| ARSH | 5.90E-20 | 1.19E-06 | 2.20E-05 | 4.12E-21 | 5.90E-20 |
| DMD | 2.01E-38 | 8.39E-15 | 3.54E-18 | 1.32E-52 | 2.01E-38 |
| *MAGEA4* | 3.11E-65 | 2.32E-22 | 7.75E-23 | 2.25E-18 | 1.48E-52 |
| *SHROOM2* | 5.41E-32 | 1.48E-11 | 6.67E-13 | 1.40E-46 | 5.41E-32 |

### The Maternal Origin of Candidate Variants of X-chromosome Genes

We identified 20 candidate X-chromosomal variants from known and novel genes affecting at least two cases, based on variant screening results (Supplementary Tables 3 and 4). There are two possibilities for the origin of these variants: *de novo* mutation during the early embryonic development of probands or during maternal oogenesis and the maternal X inheritance. For the *de novo* mutation hypothesis, we would not expect to find the variant in maternal genotypes. For the maternal X inheritance mode, we would detect heterozygotes in maternal samples.

In the seven trios with WGS data, the candidate variant NC_000023.11:g.151924064G>A; NP_001011550.1:p.(A134T) in *MAGEA4* was detected only in proband U8 (Supplementary Table 4). The Sanger Sequencing of these variants in trios indicated a maternal X-chromosome origin (Figure 2a). For the duo of U9 and 9M samples, we also observed maternal heterozygotes in the X-chromosomal variant genotype, despite the unavailability of the paternal sample. For the additional trios with WGS-only probands, the Sanger Sequencing of parental samples confirmed the maternal X inheritance, in which X hemizygotes, maternal heterozygotes, and paternal wild types were found simultaneously in all trios (Figure 2). Together, the Sanger Sequencing of four variants, which affected seven cases and parents, supported the maternal X-chromosome origin exclusively, rather than *de novo* mutation from probands’ embryogenesis or maternal germline. Future studies based on more pedigree data are needed to further evaluate the probability of *de novo* mutation during oogenesis of maternal germline or early development of probands.

**Figure 2.**
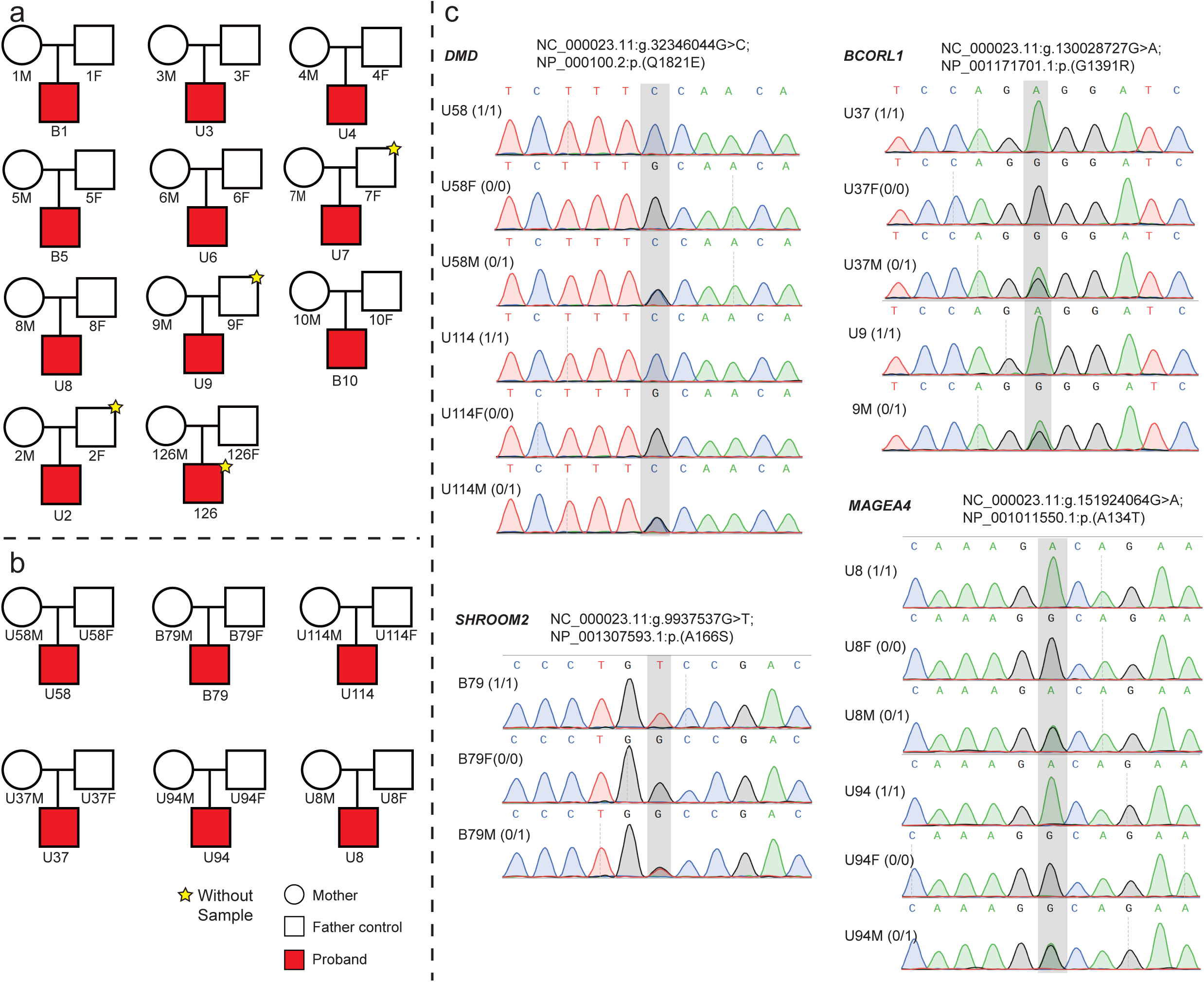
The Pedigree Information and the Chromatogram of the Sanger Sequencing Results For Cases and Their Parents. (a) The pedigree information for seven trios and four duos with WGS data. (b) The pedigree information for six trios, with WGS data only for the cases. (c) The Sanger sequencing chromatogram for candidate variants of known and novel genes. “1/1”, “0/1”, and “0/0” are genotypes. U and B represent “unilateral” and “bilateral”, respectively. “F” and “M” indicate “Father” and “Mother”, respectively. “-F” and “-R” show the results of PCR sequencing from forward primer and reverse primer, respectively.

**Figure 3.**
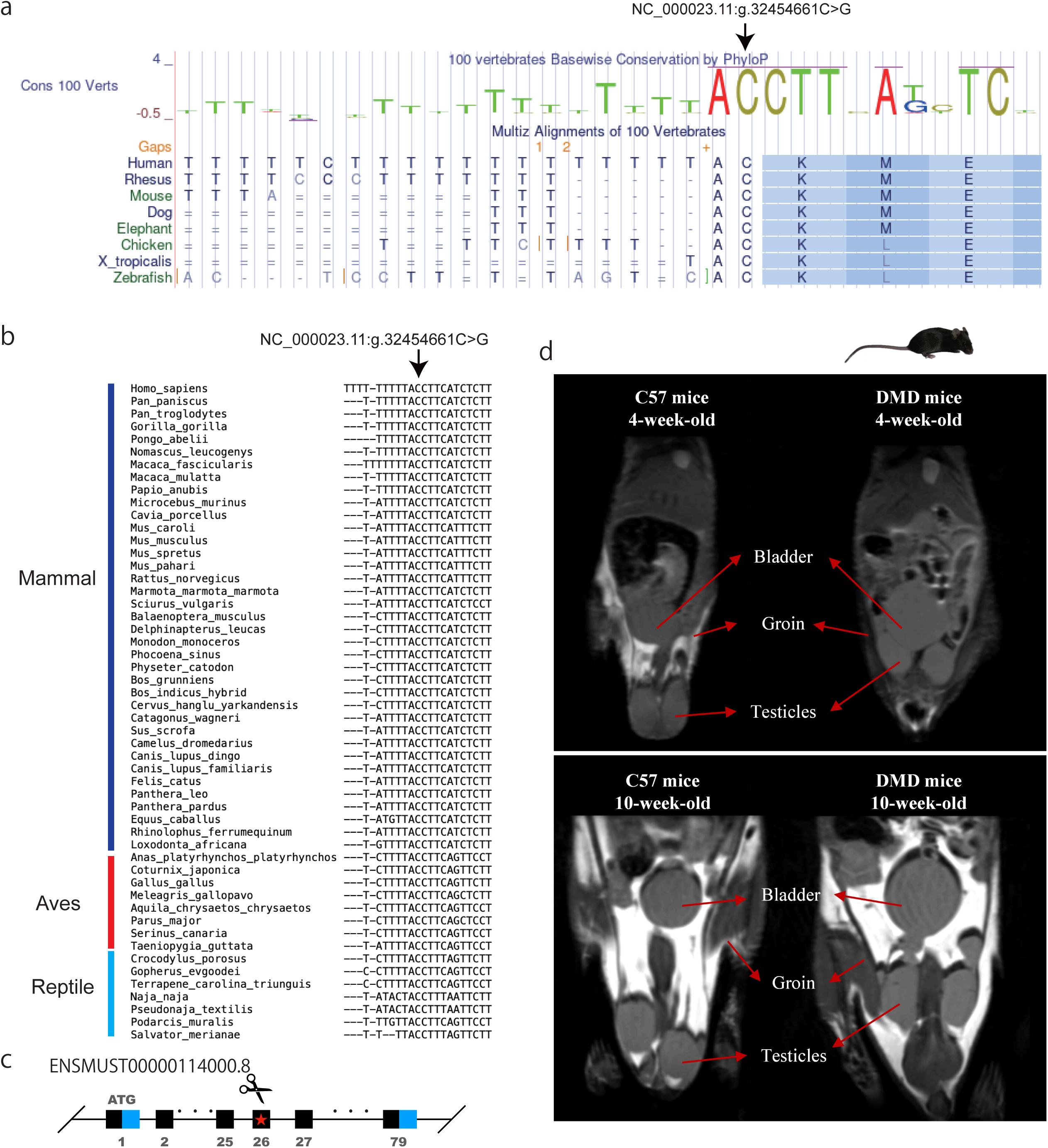
Evolutionary Conservation of the Splice Donor Variant in *DMD* (NC_000023.11:g.32454661C>G) and MRI of Transgenic and Normal Mice. (a) Conservation of the *DMD* variant (NC_000023.11:g.32454661C>G) according to the UCSC Vertebrate Alignment. (b) Conservation based on the Amniote alignment from the Ensembl database. (c) Schematic representation of a transgenic mouse with CRISPR-Cas9 editing. (d) Position of the testicles in normal C57 mice compared to *DMD* transgenic mice, as determined by MRI. Undescended testicles were observed in the *DMD* transgenic mice at both 4 and 10 weeks, with both testicles located in the inguinal region.

### No Reliable Compound Heterozygous Rare Variants Were Detected

For compound heterozygous mode in autosomes, the family-based structure can facilitate tracing of the parental origin for each rare variant. We firstly focused on two heterozygous rare variants with disruptive effects on proteins for patients of the seven complete trio families. Then, we relaxed the criteria of variant impacts to cover the scenario of one heterozygous rare variant with disruptive effect and the other with moderate impact (sequence-altering). We did not find any gene fulfilling the requirements. Finally, we focused on the scenario of potential compound heterozygous mode, in which non-pedigree probands would carry at least two heterozygous rare variants in a gene with disruptive effects. We observed two heterozygous rare variants of *RPTN* with stop-gain and frameshift impacts in four unrelated patients (Supplementary Table 5). However, the two variants had the same allele frequency in all super-populations in the gnomAD database, suggesting that they are more likely to be inherited via the linkage disequilibrium, rather than the real compound heterozygous variants.

### CRISPR-Cas9 mouse modeling of the splice donor variant in *DMD* (NC_000023.11:g.32454661C>G)

Considering the highly conserved nature of the wild-type splice donor variant in *DMD* (NC_000023.11:g.32454661C>G, represented as ‘C’ in Figure 3a-3b), we generated transgenic mice with the mutant variant (‘G’) using the CRISPR-Cas9 editing technique (Figure 3c). We examined cryptorchidism phenotypes in F2 male mice using MRI at weeks 4 and 10. In the *DMD* transgenic mice, we found undescended testicles that are different from the wild-type mice of the same age (Figure 3d). The undescended testicles were located in the inguinal region, suggesting the involvement of the *DMD* gene in the development of cryptorchidism.

## Discussion

In this study, we conducted rare variants screening for cryptorchidism in humans. Among the known genes related to cryptorchidism, the mutation burden and rigorous filtering of rare and predicted deleterious variants support the roles of *USP9Y*, *KDM6A*, *BCORL1*, and *UBA1* in the pathogenicity of cryptorchidism. *USP9Y* is one of the three genes within an azoospermia factor (AZFa) region. Previous studies have revealed that *USP9Y* showed increased transcript levels in patients suffering from cryptorchidism after a treatment with the gonadotropin-releasing hormone agonist GnRHa ^35^. Exome sequencing has revealed the role of *BCORL1* in spermatogenesis ^36^. *UBA1* has been reported in patients with Spinal Muscular Atrophy 2 and cryptorchidism ^37^. *KDM6A* is related to the Kabuki syndrome, which may involve hypospadias and cryptorchidism in males ^38^. However, most of these studies were not designed specifically for investigating cryptorchidism-associated genes and variants, so our study expands the phenotypic spectrum of these gene defects.

More importantly, we identified four novel candidate cryptorchid genes (*DMD*, *ARSH*, *MAGEA4*, and *SHROOM2*) with genome-wide significant burdens of rare mutations with both disruptive and sequence-altering impacts. The gene *MAGEA4* (melanoma antigen family A, 4) is commonly used as a marker for human spermatogonia ^39^. Based on the immunohistochemistry of *MAGEA4*, the number of spermatogonia was decreased in cryptorchid testes compared to the normal testes ^40^. Although poorly characterized, the gene *SHROOM2* was reported in patients with infertility ^41^. Notably, the findings of the pathogenic variants of *DMD* are particularly interesting, because of the established role of *DMD* in producing the protein dystrophin critical for muscle development ^42^. Clinical studies have revealed a positive correlation (OR, 2.83) between the urogenital malformation and muscular disorders, possibly due to muscular defects in the cremaster muscle or other inguinal tissues ^43^. *DMD* defects have been reported extensively in Duchenne/Becker muscular dystrophy ^44^ ^45^. Considering the dynamic process involving the muscular traction of the gubernaculum during the normal testes descendance ^46^, our study suggests that muscular abnormalities may play a major role in cryptorchidism.

As the maternal effect is higher than the paternal effect for cryptorchidism (for example, the recurrence rate is higher in maternal half-brothers than in paternal half-brothers), the possibility of maternal inheritance has long been suspected ^9^ ^47^. In this study, we revealed the genetic basis of maternal inheritance of cryptorchidism by finding the dominant role of X-chromosomal genes and their maternal origin. Indeed, based on the pedigree-based Sanger sequencing, we revealed that some X-chromosomal variants with available parental samples were inherited from maternal rather than paternal lineages. This result is consistent with the expectation of both the X-hemizygosity effect and the nature of male-specific disease. Because of X-hemizygosity in males, X-chromosomal pathogenic variants invariably affect the males, while commonly being masked by a normal allele in heterozygous females. In addition, the causative variant for a male-specific trait would affect males more heavily than females, except in case of a pleiotropic effect of the variant on both sexes. Clinically, such hereditary pattern could be utilized to screen those sex-related genetic disorders in male humans and to develop subsequent prevention and therapeutic measurements.

Together, among the total of eight known and novel genes affecting multiple unrelated patients of cryptorchidism, seven are X-chromosomal genes and one is a Y gene. The candidate variants of X-chromosomal genes follow the expectation of maternal inheritance rather than *de novo* mutation. Thus, our study revealed a predominant role of sex chromosomal, and particularly the X-chromosomal, gene defects, in causing male cryptorchidism.

## Conclusion

We identified rare pathogenic variants of four known candidate genes (*USP9Y*, *UBA1*, *BCORL1*, and *KDM6A*) and four novel candidate genes (*ARSH*, *DMD*, *MAGEA4*, and *SHROOM2*) in patients suffering from cryptorchidism. Considering the chromosomal distribution, seven out of eight genes are within the X-chromosome, and *USP9Y* is located on the Y-chromosome. This reveals a predominant role of X-chromosomal genes in cryptorchidism. The maternal origin of these X-chromosomal variants reflects the strong effect of X-hemizygosity on male-specific diseases. We successfully replicated cryptorchidism phenotypes in transgenic mice. These mice harbor a splice donor loss variant in *DMD* (NC_000023.11:g.32454661C>G), establishing a viable mouse model for future research and therapy studies of this condition.

## Materials and methods

### DNA samples, patient background, and whole-genome sequencing

DNA was extracted from peripheral whole blood of 115 patients and 19 parental controls, using a local database of variants from 2136 unaffected men for subsequent comparison. The patients were diagnosed by pediatric specialists of the West China Hospital (WCH) and West China Second University Hospital (WCSUH). The parents were carefully inquired about family history and all fathers denied the existence of cryptorchidism in the family history. All participating parents provided informed consent, and this study was formally approved by the ethics committees of WCH (Registration number: 2021389) and WCSUH (Registration number: 2021389). The WGS (150 bp paired-end) data of an insert size of 350 bp were sequenced using the DNBSEQ-T7 platform (MGI), according to the manufacturer’s protocol (Supplementary Table 1).

### Variants calling, genotyping, and annotation

Based on the high-performance computing system, we locally conducted the variant calling, genetic relationship, and population ancestry using the pipeline described previously ^48–50^. The rare variants were defined as alleles with frequency lower than 0.01 in all geographic human populations of gnomAD v3.1. Pathogenic variants were identified for known and novel genes (Supplementary note).

### Validation of variants and genotypes using the Sanger sequencing

Following the identification of candidate rare pathogenic variants, the Sanger sequencing was conducted for all probands at first to remove false positive variants due to errors in NGS sequencing or calling process. For the validated variants in probands, we further conducted the Sanger sequencing for all available parental samples, including samples from additional six trios in which only probands had WGS data. The parental genotypes were further used to evaluate the origin of variants.

### The C56BL/6J transgenic mouse modeling

For the highly conserved variant, transgenic mouse was modeled using the CRISPR-Cas9 editing technique. Briefly, the process was carried out in two main stages: the in vitro stage and the in vivo stage. In the in vitro stage, the process began with the design and construction of guide RNAs (gRNAs) specific to the target DNA sequence around variant, followed by the design and construction of the donor vector, which carried the desired genetic modification. The in vivo stage involved microinjecting the designed gRNA and donor vector into fertilized eggs, and then transplanting these embryos into surrogate mothers. This led to the birth of the F0 generation mice, which were subsequently screened to identify individuals with successful genetic modifications. Positive F0 generation mice were then bred to produce the F1 generation, and these offspring were also screened to confirm the presence of the genetic modification. The F2 mice were examined for phenotypes with MRI (Time Medical Systems, Inc, USA).

**Statements & Declarations**

## Acknowledgements

This study was supported by the fifth batch of technological innovation research projects in Chengdu (2021-YF05 -01331-SN), the Postdoctoral Research and Development Fund of West China Hospital of Sichuan University (2020HXBH087), the Short-Term Expert Fund of West China Hospital (139190032), Fund of Sichuan Provincial Department of Science and Technology (2021YFS0244), and Fund of Sichuan Provincial Department of Science and Technology (2021YFS0026). We also acknowledge the computing support from the West China Biomedical Big Data Center and the Med-X Center for Informatics of Sichuan University.

## Competing Interests

The authors declare no competing financial interests.

## Author Contributions

H.Y.X., C.Z.F., L.Z., and F.P.L. supervised this work. J.H.C., J.Z., K.Z., and H.Z.D., designed the research. J.H.C., J.Z., and GL.H analyzed data. J.H.C, Y.Y.J, and J.Z. conducted lab work. G.L.H provided the candidate rare variants screening in 2136 normal controls. J.H.C. drafted the manuscript and all authors took part in revising and improving the manuscript.

## Data Availability

The candidate variants data are listed in the supplementary tables. The variants data could be available from the corresponding author on request.

## Ethics approval

All authors declare that they have no Conflict of Interest. This study conformed with the Helsinki Declaration of 1975 (as revised in 2008) concerning Human and Animal Rights. All participating parents provided informed consent, and this study was formally approved by the ethics committees of WCH (Registration number: 2021389) and WCSUH (Registration number: 2021389).

## Consent to participate

Written informed consent was obtained from the parents.

## Notes

### Competing Interest Statement

The authors have declared no competing interest.

