## Supplementary figures and images for "Novel mutation leading to splice donor loss in a conserved site of *DMD* causes cryptorchidism"

### Supplementary Figure 1

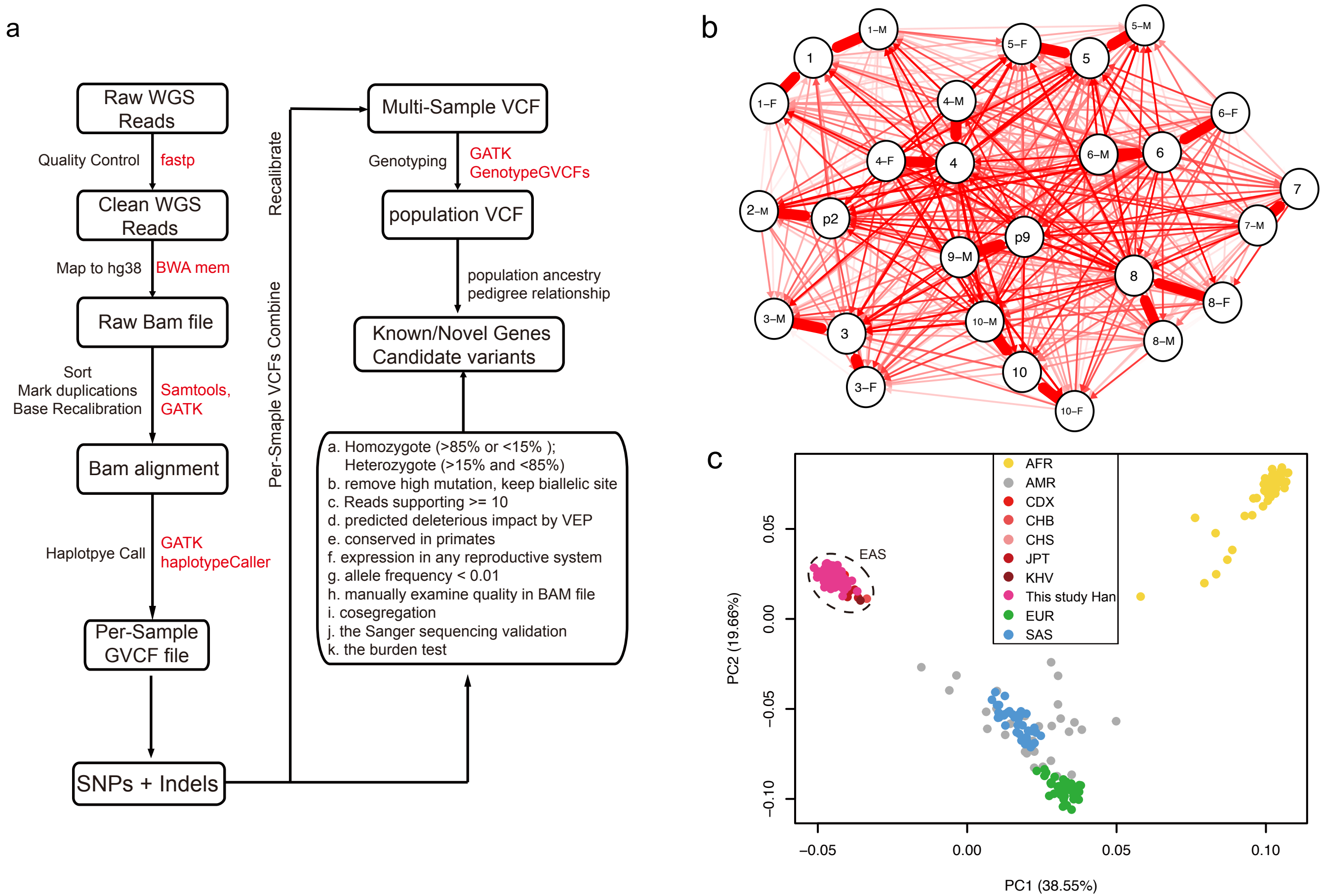

### Supplementary Figure 2

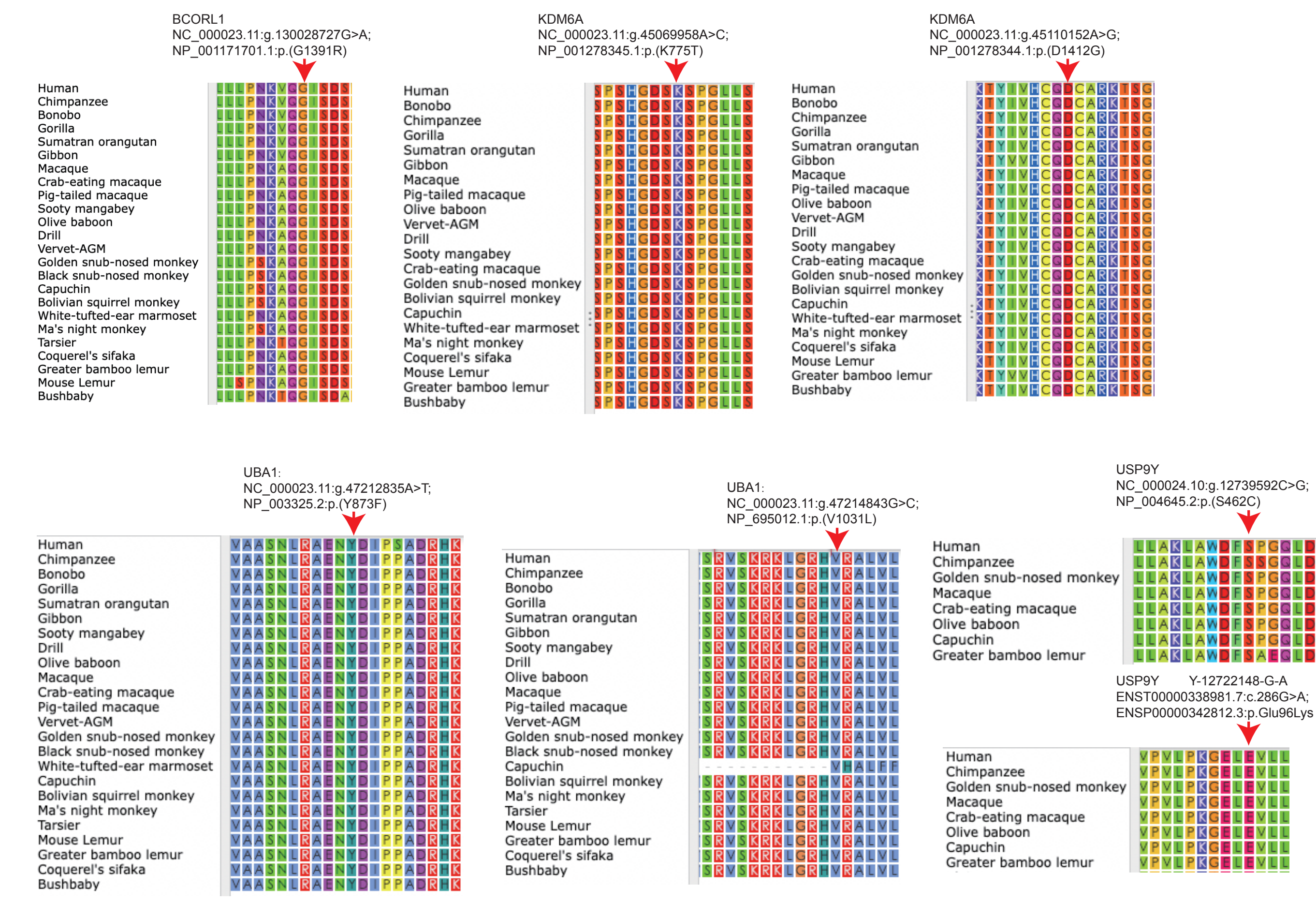

### Supplementary Figure 3

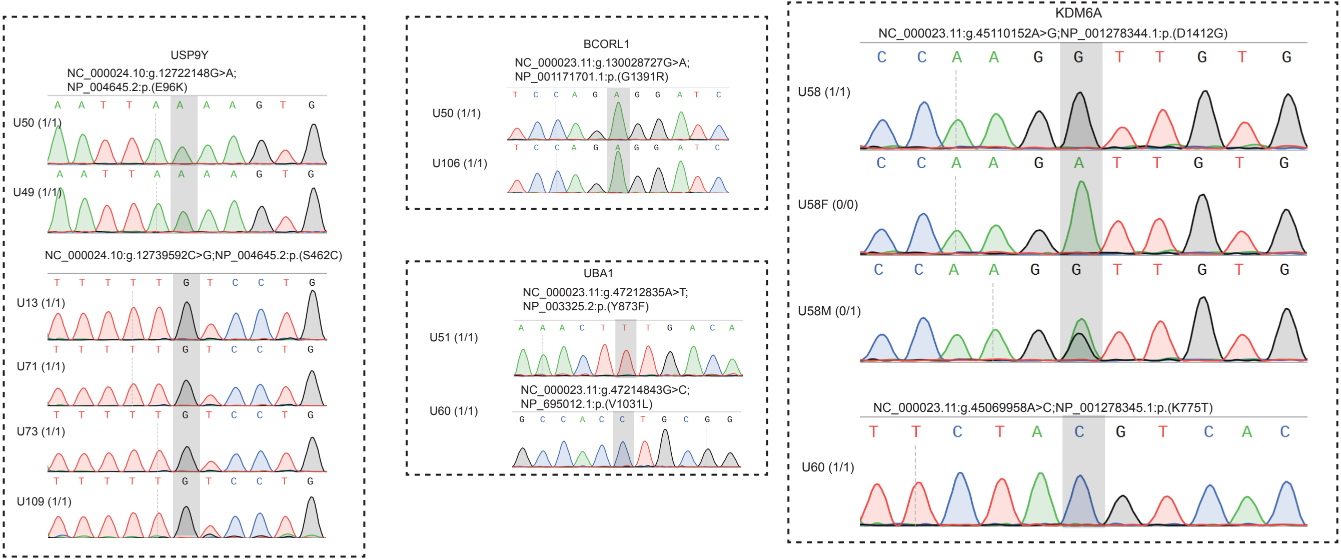

### Supplementary Figure 4

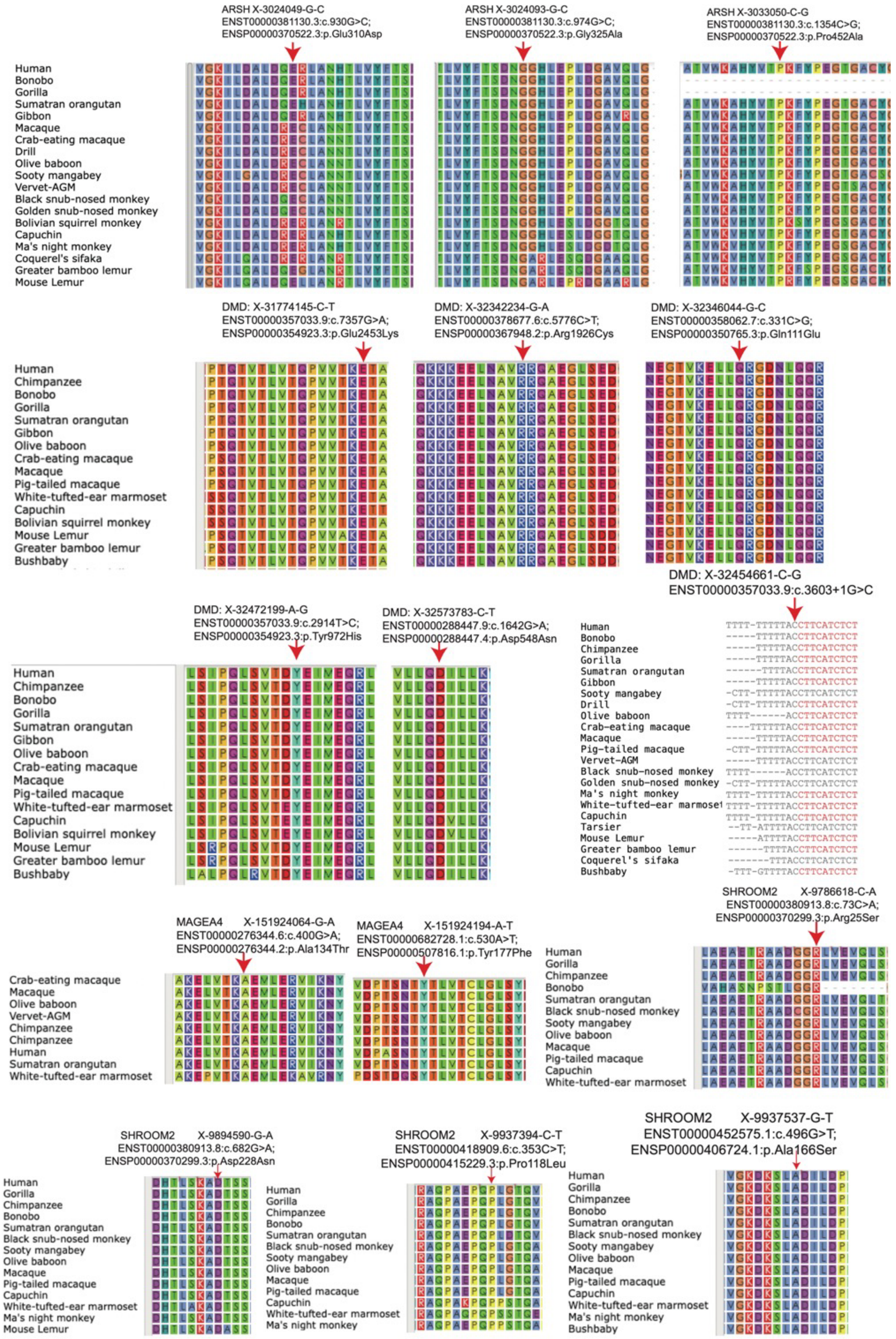

### Supplementary Figure 5

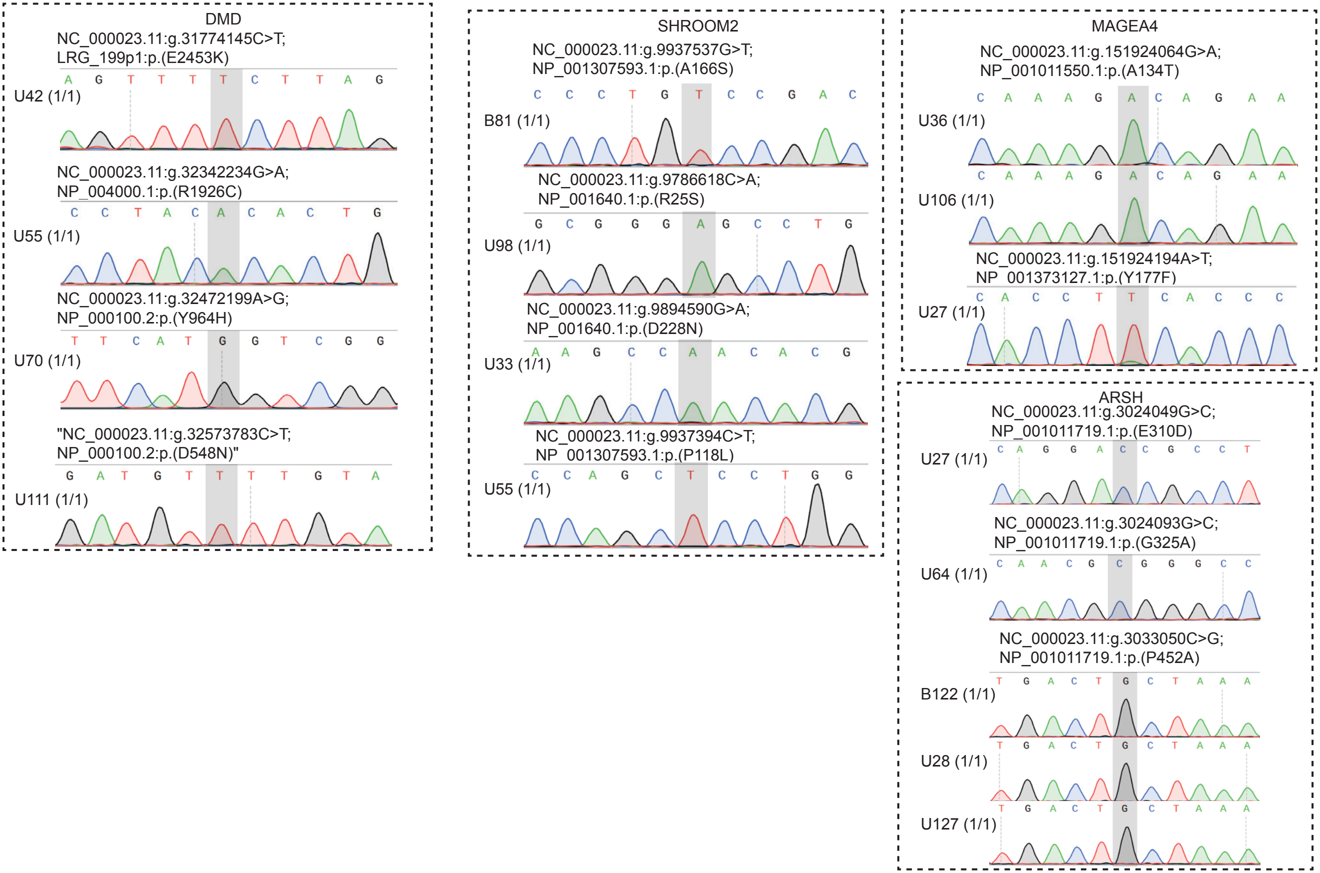
