## Supplementary table for "Novel mutation leading to splice donor loss in a conserved site of *DMD* causes cryptorchidism"

**Supplementary table 1. Anonymized samples, Phenotypes, sequencing depth and coverage. Notably, the sample ID tagged with "-F" and "-M"are fathers and mothers, respectively.**

| **Anonymized ID** | **Phenotype** | **Affected** | **Depth** | **Coverage** |
| --- | --- | --- | --- | --- |
| B1 | Bilateral | case | 32.95 | 91.38% |
| 1-F | control | control | 31.72 | 91.32% |
| 1-M | control | control | 26.93 | 90.83% |
| U2 | Unilateral | case | 25.82 | 91.44% |
| U3 | Unilateral | case | 25.72 | 91.43% |
| 3-F | control | control | 28.05 | 91.28% |
| 3-M | control | control | 26.97 | 90.61% |
| U4 | Unilateral | case | 30.72 | 91.52% |
| 4-F | control | control | 33.53 | 91.51% |
| 4-M | control | control | 38.19 | 90.95% |
| B5 | Bilateral | case | 26.51 | 91.23% |
| 5-F | control | control | 28.58 | 91.27% |
| 5-M | control | control | 26.58 | 90.60% |
| U6 | Unilateral | case | 25.16 | 91.41% |
| 6-F | control | control | 26.21 | 91.39% |
| 6-M | control | control | 25.75 | 90.83% |
| U7 | Unilateral | case | 25.7 | 91.54% |
| U8 | Unilateral | case | 27.99 | 91.24% |
| 8-F | control | control | 26.41 | 91.22% |
| 8-M | control | control | 26.96 | 90.63% |
| U9 | Unilateral | case | 24.87 | 91.41% |
| B10 | Bilateral | case | 24.68 | 91.43% |
| 10-F | control | control | 27.4 | 91.23% |
| 10-M | control | control | 26.02 | 90.60% |
| U11 | Unilateral | case | 34.58 | 91.48% |
| U13 | Unilateral | case | 33.25 | 91.47% |
| B14 | Bilateral | case | 33.45 | 91.46% |
| B15 | Bilateral | case | 36.83 | 91.51% |
| U16 | Unilateral | case | 30.32 | 91.34% |
| U17 | Unilateral | case | 32.08 | 91.35% |
| U18 | Unilateral | case | 20.33 | 91.35% |
| B19 | Bilateral | case | 32.39 | 91.47% |
| B20 | Bilateral | case | 34.41 | 91.33% |
| U22 | Unilateral | case | 35.29 | 91.39% |
| U24 | Unilateral | case | 33.06 | 91.40% |
| U25 | Unilateral | case | 37.44 | 91.48% |
| U26 | Unilateral | case | 36.17 | 91.53% |
| U27 | Unilateral | case | 37.96 | 91.48% |
| U28 | Unilateral | case | 38.3 | 91.51% |
| U29 | Unilateral | case | 36.2 | 91.50% |
| U30 | Unilateral | case | 34.9 | 91.40% |
| U31 | Unilateral | case | 31.04 | 91.45% |
| U32 | Unilateral | case | 30.06 | 91.46% |
| U33 | Unilateral | case | 37.31 | 91.53% |
| B34 | Bilateral | case | 28.25 | 91.46% |
| U35 | Unilateral | case | 28.72 | 91.37% |
| U36 | Unilateral | case | 51.55 | 91.50% |
| U37 | Unilateral | case | 42.36 | 91.41% |
| U38 | Unilateral | case | 29.11 | 91.41% |
| U40 | Unilateral | case | 26.68 | 91.35% |
| U41 | Unilateral | case | 35.46 | 91.52% |
| U42 | Unilateral | case | 41.05 | 91.45% |
| B43 | Bilateral | case | 33.86 | 91.45% |
| U44 | Unilateral | case | 30.42 | 91.49% |
| U45 | Unilateral | case | 29.09 | 91.30% |
| U46 | Unilateral | case | 47.89 | 91.47% |
| U47 | Unilateral | case | 33.31 | 91.50% |
| U48 | Unilateral | case | 38.24 | 91.49% |
| U49 | Unilateral | case | 29.85 | 91.43% |
| U50 | Unilateral | case | 19.96 | 91.34% |
| U51 | Unilateral | case | 29.69 | 91.39% |
| U52 | Unilateral | case | 31.36 | 91.39% |
| U53 | Unilateral | case | 32.05 | 91.49% |
| U55 | Unilateral | case | 30.47 | 91.45% |
| U56 | Unilateral | case | 28.49 | 91.45% |
| B57 | Bilateral | case | 26.85 | 91.34% |
| U58 | Unilateral | case | 26.27 | 91.47% |
| U59 | Unilateral | case | 34.84 | 91.46% |
| U60 | Unilateral | case | 34.05 | 91.36% |
| U61 | Unilateral | case | 39.85 | 91.42% |
| B62 | Bilateral | case | 40.95 | 91.49% |
| U63 | Unilateral | case | 29.47 | 91.32% |
| U64 | Unilateral | case | 27.48 | 91.44% |
| U65 | Unilateral | case | 36.56 | 91.37% |
| U66 | Unilateral | case | 27.72 | 91.34% |
| U67 | Unilateral | case | 31.39 | 91.46% |
| U68 | Unilateral | case | 30.62 | 91.33% |
| U69 | Unilateral | case | 39.97 | 91.44% |
| U70 | Unilateral | case | 34.13 | 91.48% |
| U71 | Unilateral | case | 27.24 | 91.40% |
| B72 | Bilateral | case | 27.81 | 91.43% |
| U73 | Unilateral | case | 38.28 | 91.52% |
| B74 | Bilateral | case | 42.16 | 91.55% |
| U75 | Unilateral | case | 31.55 | 91.45% |
| U76 | Unilateral | case | 47.11 | 91.44% |
| B77 | Bilateral | case | 32.4 | 91.44% |
| U78 | Unilateral | case | 19.38 | 91.31% |
| B79 | Bilateral | case | 31.46 | 91.47% |
| U80 | Unilateral | case | 28.76 | 91.30% |
| B81 | Bilateral | case | 48.65 | 91.49% |
| U83 | Unilateral | case | 35.77 | 91.51% |
| U84 | Unilateral | case | 44.79 | 91.45% |
| U85 | Unilateral | case | 31.51 | 91.42% |
| U86 | Unilateral | case | 28.51 | 91.48% |
| U87 | Unilateral | case | 36.93 | 91.51% |
| U88 | Unilateral | case | 32.93 | 91.26% |
| U89 | Unilateral | case | 46.27 | 91.43% |
| B90 | Bilateral | case | 26.72 | 91.35% |
| U91 | Unilateral | case | 34.93 | 91.44% |
| U92 | Unilateral | case | 27.37 | 91.49% |
| U93 | Unilateral | case | 42.03 | 91.50% |
| U94 | Unilateral | case | 38.04 | 91.49% |
| U96 | Unilateral | case | 28.49 | 91.31% |
| U97 | Unilateral | case | 37.41 | 91.46% |
| U98 | Unilateral | case | 38.3 | 91.66% |
| U99 | Unilateral | case | 28.2 | 91.46% |
| U100 | Unilateral | case | 19.38 | 91.30% |
| U101 | Unilateral | case | 34.71 | 91.38% |
| B102 | Bilateral | case | 31.64 | 91.46% |
| U103 | Unilateral | case | 30.24 | 91.46% |
| U104 | Unilateral | case | 45.02 | 91.48% |
| U106 | Unilateral | case | 34.44 | 91.37% |
| U107 | Unilateral | case | 37.31 | 91.48% |
| U108 | Unilateral | case | 27.44 | 91.40% |
| U109 | Unilateral | case | 45.43 | 91.42% |
| U110 | Unilateral | case | 27.94 | 91.42% |
| U111 | Unilateral | case | 30.74 | 91.32% |
| U112 | Unilateral | case | 30.41 | 91.45% |
| U113 | Unilateral | case | 28.88 | 91.48% |
| U114 | Unilateral | case | 36.4 | 91.40% |
| U115 | Unilateral | case | 35.7 | 91.48% |
| U116 | Unilateral | case | 35.27 | 91.45% |
| U119 | Unilateral | case | 29.12 | 91.48% |
| U120 | Unilateral | case | 31.75 | 91.43% |
| B121 | Bilateral | case | 34.01 | 91.46% |
| B122 | Bilateral | case | 43.85 | 91.47% |
| U123 | Unilateral | case | 37.39 | 91.42% |
| U127 | Unilateral | case | 29.43 | 91.39% |
| 126-F | control | control | 25.17 | 91.45% |
| 126-M | control | control | 26.77 | 90.82% |
| 2-M | control | control | 27.43 | 90.87% |
| 7-M | control | control | 27.77 | 90.75% |
| 9-M | control | control | 24.91 | 90.76% |
| B130 | Bilateral | case | 34.82 | 91.32% |

**Supplementary table 2. The known gene pool from literature review and HPO database**

| **Gene** | **Chr** | **HPOlist** | **Gene annotation(NCBI)** | **Reference** |
| --- | --- | --- | --- | --- |
| ACADVL | 17 | *NO* | Acyl-CoA dehydrogenase very long chain.The protein encoded by this gene is targeted to the inner mitochondrial membrane where it catalyzes the first step of the mitochondrial fatty acid beta-oxidation pathway. This acyl-Coenzyme A dehydrogenase is specific to long-chain and very-long-chain fatty acids. | [(Bo, Awano et al. 2021)](applewebdata://FA9EE35C-7824-4A91-9A1E-4D2B671FD335/#_ENREF_14) |
| ADAT3 | 19 | *Yes* | Adenosine deaminase tRNA specific 3.This gene encodes a subunit of a tRNA-specific adenosine deaminase. This heterodimeric enzyme converts adenosine to inosine in the tRNA anticodon. | [(Thomas, Lewis et al. 2019)](applewebdata://FA9EE35C-7824-4A91-9A1E-4D2B671FD335/#_ENREF_71) |
| AMH | 19 | *Yes* | Anti-Mullerian hormone.This gene encodes a secreted ligand of the TGF-beta (transforming growth factor-beta) superfamily of proteins. Ligands of this family bind various TGF-beta receptors leading to recruitment and activation of SMAD family transcription factors that regulate gene expression. The encoded preproprotein is proteolytically processed to generate N- and C-terminal cleavage products that homodimerize and associate to form a biologically active noncovalent complex. This complex binds to the anti-Mullerian hormone receptor type 2 and causes the regression of Mullerian ducts in the male embryo that would otherwise differentiate into the uterus and fallopian tubes. This protein also plays a role in Leydig cell differentiation and function and follicular development in adult females. | (Acero, Moreno et al. 2019, Liu, Wang et al. 2022), |
| AMHR2 | 12 | *Yes* | Anti-Mullerian hormone receptor type 2.This gene encodes the receptor for the anti-Mullerian hormone (AMH) which, in addition to testosterone, results in male sex differentiation. AMH and testosterone are produced in the testes by different cells and have different effects. Testosterone promotes the development of male genitalia while the binding of AMH to the encoded receptor prevents the development of the mullerian ducts into uterus and Fallopian tubes. | (Acero, Moreno et al. 2019, Liu, Wang et al. 2022), |
| ANKRD11 | 16 | *Yes* | Ankyrin repeat domain 11.The encoded protein inhibits ligand-dependent activation of transcription. | [(Sayed, Abdel-Hamid et al. 2020)](applewebdata://FA9EE35C-7824-4A91-9A1E-4D2B671FD335/#_ENREF_61) |
| AR | X | *Yes* | The AR gene provides instructions for making a protein called an androgen receptor. | [(Kaftanovskaya, Huang et al. 2012)](applewebdata://FA9EE35C-7824-4A91-9A1E-4D2B671FD335/#_ENREF_34) |
| ATRX | X | Yes | ATRX chromatin remodeler.The protein encoded by this gene contains an ATPase/helicase domain, and thus it belongs to the SWI/SNF family of chromatin remodeling proteins. This protein is found to undergo cell cycle-dependent phosphorylation, which regulates its nuclear matrix and chromatin association, and suggests its involvement in the gene regulation at interphase and chromosomal segregation in mitosis. Mutations in this gene are associated with X-linked syndromes exhibiting cognitive disabilities as well as alpha-thalassemia (ATRX) syndrome. | [(Villard, Fontès et al. 2000)](applewebdata://FA9EE35C-7824-4A91-9A1E-4D2B671FD335/#_ENREF_77) |
| CHD3 | 17 | *NO* | Chromodomain helicase DNA binding protein 3.This gene encodes a member of the CHD family of proteins which are characterized by the presence of chromo (chromatin organization modifier) domains and SNF2-related helicase/ATPase domains. This protein is one of the components of a histone deacetylase complex referred to as the Mi-2/NuRD complex which participates in the remodeling of chromatin by deacetylating histones. | [(Coursimault, Lecoquierre et al. 2021)](applewebdata://FA9EE35C-7824-4A91-9A1E-4D2B671FD335/#_ENREF_19) |
| COL3A1 | 2 | *Yes* | Collagen type III alpha 1 chain.This gene encodes the pro-alpha1 chains of type III collagen, a fibrillar collagen that is found in extensible connective tissues such as skin, lung, uterus, intestine and the vascular system, frequently in association with type I collagen. | [(Park, Shin et al. 2017)](applewebdata://FA9EE35C-7824-4A91-9A1E-4D2B671FD335/#_ENREF_48) |
| CYP21A2 | 6 | *NO* | This gene encodes a member of the cytochrome P450 superfamily of enzymes. The cytochrome P450 proteins are monooxygenases which catalyze many reactions involved in drug metabolism and synthesis of cholesterol, steroids and other lipids. This protein localizes to the endoplasmic reticulum and hydroxylates steroids at the 21 position. Its activity is required for the synthesis of steroid hormones including cortisol and aldosterone. | [(Umino, Kitamura et al. 2019)](applewebdata://FA9EE35C-7824-4A91-9A1E-4D2B671FD335/#_ENREF_74) |
| DHDDS | 1 | Yes | Dehydrodolichyl diphosphate synthase subunit.The protein encoded by this gene catalyzes cis-prenyl chain elongation to produce the polyprenyl backbone of dolichol, a glycosyl carrier lipid required for the biosynthesis of several classes of glycoproteins. | [(Sabry, Vuillaumier-Barrot et al. 2016)](applewebdata://FA9EE35C-7824-4A91-9A1E-4D2B671FD335/#_ENREF_59) |
| EED | 11 | *Yes* | Embryonic ectoderm development.This gene encodes a member of the Polycomb-group (PcG) family. PcG family members form multimeric protein complexes, which are involved in maintaining the transcriptional repressive state of genes over successive cell generations. This protein interacts with enhancer of zeste 2, the cytoplasmic tail of integrin beta7, immunodeficiency virus type 1 (HIV-1) MA protein, and histone deacetylase proteins. This protein mediates repression of gene activity through histone deacetylation, and may act as a specific regulator of integrin function. | [(Griffiths, Loveday et al. 2019)](applewebdata://FA9EE35C-7824-4A91-9A1E-4D2B671FD335/#_ENREF_28) |
| EHMT1 | 9 | *Yes* | Euchromatic histone lysine methyltransferase 1.The protein encoded by this gene is a histone methyltransferase that methylates the lysine-9 position of histone H3. This action marks the genomic region packaged with these methylated histones for transcriptional repression. This protein may be involved in the silencing of MYC- and E2F-responsive genes and therefore could play a role in the G0/G1 cell cycle transition. | [(Torga, Hodax et al. 2018)](applewebdata://FA9EE35C-7824-4A91-9A1E-4D2B671FD335/#_ENREF_73) |
| EIF2S3 | X | *Yes* | Eukaryotic translation initiation factor 2 subunit gamma.The protein encoded by this gene is the largest subunit of a heterotrimeric GTP-binding protein involved in the recruitment of methionyl-tRNA(i) to the 40 S ribosomal subunit. | [(Mori, Kumada et al. 2021)](applewebdata://FA9EE35C-7824-4A91-9A1E-4D2B671FD335/#_ENREF_41) |
| EZH2 | 7 | *Yes* | Enhancer of zeste 2 polycomb repressive complex 2 subunit. This gene encodes a member of the Polycomb-group (PcG) family. PcG family members form multimeric protein complexes, which are involved in maintaining the transcriptional repressive state of genes over successive cell generations. This protein associates with the embryonic ectoderm development protein, the VAV1 oncoprotein, and the X-linked nuclear protein. This protein may play a role in the hematopoietic and central nervous systems. | [(Griffiths, Loveday et al. 2019)](applewebdata://FA9EE35C-7824-4A91-9A1E-4D2B671FD335/#_ENREF_28) |
| FGD1 | X | *Yes* | FYVE, RhoGEF and PH domain containing 1.The encoded protein specifically binds to the Rho family GTPase Cdc42Hs and can stimulate the GDP-GTP exchange of the isoprenylated form of Cdc42Hs. It also stimulates the mitogen activated protein kinase cascade leading to c-Jun kinase SAPK/JNK1 activation. | [(Jia, Ma et al. 2021)](applewebdata://FA9EE35C-7824-4A91-9A1E-4D2B671FD335/#_ENREF_32) |
| GREAT | 13 | NO | Relaxin family peptide receptor 2.This gene encodes a member of the GPCR (G protein-coupled, 7-transmembrane receptor) family. | (Gorlov, Kamat et al. 2002), ANKRD11 (Sirmaci, Spiliopoulos et al. 2011) |
| HDAC2 | 6 | *NO* | Histone deacetylase 2.This protein forms transcriptional repressor complexes by associating with many different proteins, including YY1, a mammalian zinc-finger transcription factor. Thus, it plays an important role in transcriptional regulation, cell cycle progression and developmental events. | [(Wagner, Hillman et al. 2019)](applewebdata://FA9EE35C-7824-4A91-9A1E-4D2B671FD335/#_ENREF_79) |
| HSPA9 | 5 | *NO* | Heat shock protein family A (Hsp70) member 9.This gene encodes a member of the heat shock protein 70 gene family. The encoded protein is primarily localized to the mitochondria but is also found in the endoplasmic reticulum, plasma membrane and cytoplasmic vesicles. This protein is a heat-shock cognate protein. This protein plays a role in cell proliferation, stress response and maintenance of the mitochondria. | [(Younger, Vetrini et al. 2020)](applewebdata://FA9EE35C-7824-4A91-9A1E-4D2B671FD335/#_ENREF_85) |
| IER3IP1 | 18 | *Yes* | Immediate early response 3 interacting protein 1.This gene encodes a small protein that is localized to the endoplasmic reticulum (ER) and may play a role in the ER stress response by mediating cell differentiation and apoptosis. Transcription of this gene is regulated by tumor necrosis factor alpha and specificity protein 1 (Sp1). | [(Rjiba, Soyah et al. 2021)](applewebdata://FA9EE35C-7824-4A91-9A1E-4D2B671FD335/#_ENREF_55) |
| INSL3 | *19* | *Yes* | Insulin-like 3 is a protein hormone produced by Leydig cells | (Adham, Emmen et al. 2000, Tomboc, Lee et al. 2000) |
| KAT6A | 8 | *Yes* | Lysine acetyltransferase 6A.The protein is composed of a nuclear localization domain, a double C2H2 zinc finger domain that binds to acetylated histone tails, a histone acetyl-transferase domain, a glutamate/aspartate-rich region, and a serine- and methionine-rich transactivation domain. It is part of a complex that acetylates lysine-9 residues in histone 3, and in addition, it acts as a co-activator for several transcription factors. | [(Jiang, Yang et al. 2021), (Urreizti, Lopez-Martin et al. 2020)](applewebdata://FA9EE35C-7824-4A91-9A1E-4D2B671FD335/#_ENREF_33) |
| KISS1R | 19 | *Yes* | KISS1 receptor.The protein encoded by this gene is a galanin-like G protein-coupled receptor that binds metastin, a peptide encoded by the metastasis suppressor gene KISS1. The tissue distribution of the expressed gene suggests that it is involved in the regulation of endocrine function, and this is supported by the finding that this gene appears to play a role in the onset of puberty. | [(Shahab, Lippincott et al. 2018)](applewebdata://FA9EE35C-7824-4A91-9A1E-4D2B671FD335/#_ENREF_64) |
| KLHL40 | 3 | *NO* | Kelch like family member 40.This gene encodes a protein containing a BACK domain, a BTB/POZ domain, and 5 Kelch repeats, however, its exact function is not known. The gene and the multi-domain protein structure are conserved across different taxa, including primates, rodents, chicken and zebrafish. | [(Yi, Zhang et al. 2021)](applewebdata://FA9EE35C-7824-4A91-9A1E-4D2B671FD335/#_ENREF_84) |
| KRAS | 12 | Yes | KRAS proto-oncogene, GTPase.This gene, a Kirsten ras oncogene homolog from the mammalian ras gene family, encodes a protein that is a member of the small GTPase superfamily. A single amino acid substitution is responsible for an activating mutation. The transforming protein that results is implicated in various malignancies, including lung adenocarcinoma, mucinous adenoma, ductal carcinoma of the pancreas and colorectal carcinoma. | [(Yao, Yu et al. 2018)](applewebdata://FA9EE35C-7824-4A91-9A1E-4D2B671FD335/#_ENREF_83) |
| LHCGR | 2 | *NO* | Luteinizing hormone/choriogonadotropin receptor.This gene encodes the receptor for both luteinizing hormone and choriogonadotropin. This receptor belongs to the G-protein coupled receptor 1 family, and its activity is mediated by G proteins which activate adenylate cyclase. | [(Jia, Ma et al. 2021)](applewebdata://FA9EE35C-7824-4A91-9A1E-4D2B671FD335/#_ENREF_32) |
| LSM1 | 8 | *NO* | This gene encodes a member of the LSm family of RNA-binding proteins. LSm proteins form stable heteromers that bind specifically to the 3'-terminal oligo(U) tract of U6 snRNA and may play a role in pre-mRNA splicing by mediating U4/U6 snRNP formation. | [(Okur, LeDuc et al. 2019)](applewebdata://FA9EE35C-7824-4A91-9A1E-4D2B671FD335/#_ENREF_47) |
| MBTPS2 | X | *Yes* | Membrane bound transcription factor peptidase, site 2.This gene encodes a intramembrane zinc metalloprotease, which functions in the signal protein activation involved in sterol control of transcription and the ER stress response. | [(Strong, March et al. 2022)](applewebdata://FA9EE35C-7824-4A91-9A1E-4D2B671FD335/#_ENREF_68) |
| MTOR | 1 | Yes | Mechanistic target of rapamycin kinase.The protein encoded by this gene belongs to a family of phosphatidylinositol kinase-related kinases. These kinases mediate cellular responses to stresses such as DNA damage and nutrient deprivation. This kinase is a component of two distinct complexes, mTORC1, which controls protein synthesis, cell growth and proliferation, and mTORC2, which is a regulator of the actin cytoskeleton, and promotes cell survival and cell cycle progression. This protein acts as the target for the cell-cycle arrest and immunosuppressive effects of the FKBP12-rapamycin complex. Inhibitors of mTOR are used in organ transplants as immunosuppressants, and are being evaluated for their therapeutic potential in SARS-CoV-2 infections. | [(Mroske, Rasmussen et al. 2015)](applewebdata://FA9EE35C-7824-4A91-9A1E-4D2B671FD335/#_ENREF_43) |
| MYRF | 11 | *Yes* | Myelin regulatory factor.This gene encodes a transcription factor that is required for central nervous system myelination and may regulate oligodendrocyte differentiation. It is thought to act by increasing the expression of genes that effect myelin production but may also directly promote myelin gene expression. | [(Rossetti, Glinton et al. 2019) (Pinz, Pyle et al. 2018)](applewebdata://FA9EE35C-7824-4A91-9A1E-4D2B671FD335/#_ENREF_57) |
| NAA10 | X | Yes | N-alpha-acetyltransferase 10, NatA catalytic subunit.N-alpha-acetylation is among the most common post-translational protein modifications in eukaryotic cells. This process involves the transfer of an acetyl group from acetyl-coenzyme A to the alpha-amino group on a nascent polypeptide and is essential for normal cell function. This gene encodes an N-terminal acetyltransferase that functions as the catalytic subunit of the major amino-terminal acetyltransferase A complex. | [(Rope, Wang et al. 2011)](applewebdata://FA9EE35C-7824-4A91-9A1E-4D2B671FD335/#_ENREF_56) |
| NONO | X | *Yes* | Non-POU domain containing octamer binding.This gene encodes an RNA-binding protein which plays various roles in the nucleus, including transcriptional regulation and RNA splicing. A rearrangement between this gene and the transcription factor E3 gene has been observed in papillary renal cell carcinoma. | [(Carlston, Bleyl et al. 2019),](applewebdata://FA9EE35C-7824-4A91-9A1E-4D2B671FD335/#_ENREF_16) |
| NR2F2 | 15 | *NO* | This gene encodes a member of the steroid thyroid hormone superfamily of nuclear receptors. The encoded protein is a ligand inducible transcription factor that is involved in the regulation of many different genes. Alternate splicing results in multiple transcript variants | [(Arsov, Kelecic et al. 2021)](applewebdata://FA9EE35C-7824-4A91-9A1E-4D2B671FD335/#_ENREF_9) |
| NR5A1 | 9 | *Yes* | Nuclear receptor subfamily 5 group A member 1.The protein encoded by this gene is a transcriptional activator involved in sex determination. The encoded protein binds DNA as a monomer. | [(Askari, Rastari et al. 2020)](applewebdata://FA9EE35C-7824-4A91-9A1E-4D2B671FD335/#_ENREF_10) |
| NSD2 | 4 | Yes | Nuclear receptor binding SET domain protein 2.This gene encodes a protein that contains four domains present in other developmental proteins: a PWWP domain, an HMG box, a SET domain, and a PHD-type zinc finger. It is expressed ubiquitously in early development. Wolf-Hirschhorn syndrome (WHS) is a malformation syndrome associated with a hemizygous deletion of the distal short arm of chromosome 4. This gene maps to the 165 kb WHS critical region and has also been involved in the chromosomal translocation t(4;14)(p16.3;q32.3) in multiple myelomas. | [(Wiel, Bruno et al. 2022)](applewebdata://FA9EE35C-7824-4A91-9A1E-4D2B671FD335/#_ENREF_82) |
| OCRL | X | *Yes* | This gene encodes an inositol polyphosphate 5-phosphatase. This protein is involved in regulating membrane trafficking and is located in numerous subcellular locations including the trans-Golgi network, clathrin-coated vesicles and, endosomes and the plasma membrane. This protein may also play a role in primary cilium formation. | [(Zhou, Wang et al. 2022)](applewebdata://FA9EE35C-7824-4A91-9A1E-4D2B671FD335/#_ENREF_88) |
| OPHN1 | X | *Yes* | Oligophrenin 1.This gene encodes a Rho-GTPase-activating protein that promotes GTP hydrolysis of Rho subfamily members. Rho proteins are important mediators of intracellular signal transduction, which affects cell migration and cell morphogenesis. | [(Schwartz, Wojcik et al. 2019)](applewebdata://FA9EE35C-7824-4A91-9A1E-4D2B671FD335/#_ENREF_63) |
| P450c17 | 10 | NO | Cytochrome P450 family 17 subfamily A member 1.This gene encodes a member of the cytochrome P450 superfamily of enzymes. The cytochrome P450 proteins are monooxygenases which catalyze many reactions involved in drug metabolism and synthesis of cholesterol, steroids and other lipids. | [(Koika, Armeni et al. 2016)](applewebdata://FA9EE35C-7824-4A91-9A1E-4D2B671FD335/#_ENREF_38) |
| PBX1 | 1 | Yes | PBX homeobox 1.This gene encodes a nuclear protein that belongs to the PBX homeobox family of transcriptional factors. Studies in mice suggest that this gene may be involved in the regulation of osteogenesis and required for skeletal patterning and programming. A chromosomal translocation, t(1;19) involving this gene and TCF3/E2A gene, is associated with pre-B-cell acute lymphoblastic leukemia. The resulting fusion protein, in which the DNA binding domain of E2A is replaced by the DNA binding domain of this protein, transforms cells by constitutively activating transcription of genes regulated by the PBX protein family. | [(Riedhammer, Siegel et al. 2017)](applewebdata://FA9EE35C-7824-4A91-9A1E-4D2B671FD335/#_ENREF_53) |
| PIGN | 18 | Yes | Phosphatidylinositol glycan anchor biosynthesis class N.This gene encodes a protein that is involved in glycosylphosphatidylinositol (GPI)-anchor biosynthesis. The GPI-anchor is a glycolipid found on many blood cells and serves to anchor proteins to the cell surface. This protein is expressed in the endoplasmic reticulum and transfers phosphoethanolamine (EtNP) to the first mannose of the GPI anchor. | [(Couser, Masood et al. 2015)](applewebdata://FA9EE35C-7824-4A91-9A1E-4D2B671FD335/#_ENREF_20) |
| POR | 7 | Yes | Cytochrome p450 oxidoreductase.This gene encodes an endoplasmic reticulum membrane oxidoreductase that is essential for multiple metabolic processes, including reactions catalyzed by cytochrome P450 proteins for metabolism of steroid hormones, drugs and xenobiotics. The encoded protein has a flavin adenine dinucleotide (FAD)-binding domain and a flavodoxin-like domain which bind two cofactors, FAD and FMN, that allow it to donate electrons directly from NADPH to all microsomal P450 enzymes. | [(Sánchez-Garvín, Albaladejo et al. 2013)](applewebdata://FA9EE35C-7824-4A91-9A1E-4D2B671FD335/#_ENREF_60) |
| PTCH1 | 9 | *Yes* | Patched 1.This gene encodes a member of the patched family of proteins and a component of the hedgehog signaling pathway. Hedgehog signaling is important in embryonic development and tumorigenesis. The encoded protein is the receptor for the secreted hedgehog ligands, which include sonic hedgehog, indian hedgehog and desert hedgehog. Following binding by one of the hedgehog ligands, the encoded protein is trafficked away from the primary cilium, relieving inhibition of the G-protein-coupled receptor smoothened, which results in activation of downstream signaling. | [(Barraud, Delemer et al. 2021)](applewebdata://FA9EE35C-7824-4A91-9A1E-4D2B671FD335/#_ENREF_12) |
| PTPN11 | 12 | *Yes* | Protein tyrosine phosphatase non-receptor type 11.PTPs are known to be signaling molecules that regulate a variety of cellular processes including cell growth, differentiation, mitotic cycle, and oncogenic transformation.This PTP is widely expressed in most tissues and plays a regulatory role in various cell signaling events that are important for a diversity of cell functions, such as mitogenic activation, metabolic control, transcription regulation, and cell migration | [(Hu, Chen et al. 2021)](applewebdata://FA9EE35C-7824-4A91-9A1E-4D2B671FD335/#_ENREF_29) |
| RAF1 | 3 | *Yes* | Raf-1 proto-oncogene, serine/threonine kinase.This gene is the cellular homolog of viral raf gene (v-raf). The encoded protein is a MAP kinase kinase kinase (MAP3K), which functions downstream of the Ras family of membrane associated GTPases to which it binds directly. Once activated, the cellular RAF1 protein can phosphorylate to activate the dual specificity protein kinases MEK1 and MEK2, which in turn phosphorylate to activate the serine/threonine specific protein kinases, ERK1 and ERK2. Activated ERKs are pleiotropic effectors of cell physiology and play an important role in the control of gene expression involved in the cell division cycle, apoptosis, cell differentiation and cell migration. | [(Kohn, Lopategui et al. 2019)](applewebdata://FA9EE35C-7824-4A91-9A1E-4D2B671FD335/#_ENREF_37) |
| RFT1 | 3 | *Yes* | RFT1 homolog.This gene encodes an enzyme which catalyzes the translocation of the Man(5)GlcNAc (2)-PP-Dol intermediate from the cytoplasmic to the luminal side of the endoplasmic reticulum membrane in the pathway for the N-glycosylation of proteins. | [(Abiramalatha, Arunachal et al. 2019)](applewebdata://FA9EE35C-7824-4A91-9A1E-4D2B671FD335/#_ENREF_2) |
| RIT1 | 1 | *Yes* | Ras like without CAAX 1.This gene encodes a member of a subfamily of Ras-related GTPases. The encoded protein is involved in regulating p38 MAPK-dependent signaling cascades related to cellular stress. This protein also cooperates with nerve growth factor to promote neuronal development and regeneration. | [(Nemcikova, Vejvalkova et al. 2016)](applewebdata://FA9EE35C-7824-4A91-9A1E-4D2B671FD335/#_ENREF_44) |
| RRAS2 | 11 | *Yes* | The encoded protein associates with the plasma membrane and may function as a signal transducer and may play an important role in activating signal transduction pathways that control cell proliferation. | [(Weinstock and Sadler 2022)](applewebdata://FA9EE35C-7824-4A91-9A1E-4D2B671FD335/#_ENREF_81) |
| RXFP2 | *13* | *NO* | Receptor for relaxin. The activity of this receptor is mediated by G proteins leading to stimulation of adenylate cyclase and an increase of cAMP. | [(Ayers, Kumar et al. 2019)](applewebdata://FA9EE35C-7824-4A91-9A1E-4D2B671FD335/#_ENREF_11) |
| SOX2 | 3 | *Yes* | a transcription factor that is essential for maintaining self-renewal, or pluripotency, of undifferentiated embryonic stem cells. | (Errichiello, Gorgone et al. 2018, Wang, Fan et al. 2021) |
| SOX3 | X | *Yes* | involved in the regulation of embryonic brain development and in determination of cell fate. | [(Du, Wang et al. 2022)](applewebdata://FA9EE35C-7824-4A91-9A1E-4D2B671FD335/#_ENREF_22) |
| SPRY4 | 5 | *Yes* | Sprouty RTK signaling antagonist 4.This gene encodes a member of a family of cysteine- and proline-rich proteins. The encoded protein is an inhibitor of the receptor-transduced mitogen-activated protein kinase (MAPK) signaling pathway. Activity of this protein impairs the formation of active GTP-RAS. Nucleotide variation in this gene has been associated with hypogonadotropic hypogonadism 17 with or without anosmia. | [(Indirli, Cangiano et al. 2019)](applewebdata://FA9EE35C-7824-4A91-9A1E-4D2B671FD335/#_ENREF_30) |
| TASP1 | 20 | Yes | Taspase 1.This gene encodes an endopeptidase that cleaves specific substrates following aspartate residues. The encoded protein undergoes posttranslational autoproteolytic processing to generate alpha and beta subunits, which reassemble into the active alpha2-beta2 heterotetramer. It is required to cleave MLL, a protein required for the maintenance of HOX gene expression, and TFIIA, a basal transcription factor. | [(Suleiman, Riedhammer et al. 2019)](applewebdata://FA9EE35C-7824-4A91-9A1E-4D2B671FD335/#_ENREF_69) |
| TBL1XR1 | 3 | *Yes* | TBL1X/Y related 1.This gene is a member of the WD40 repeat-containing gene family and shares sequence similarity with transducin (beta)-like 1X-linked (TBL1X). The protein encoded by this gene is thought to be a component of both nuclear receptor corepressor (N-CoR) and histone deacetylase 3 (HDAC 3) complexes, and is required for transcriptional activation by a variety of transcription factors. Mutations in these gene have been associated with some autism spectrum disorders, and one finding suggests that haploinsufficiency of this gene may be a cause of intellectual disability with dysmorphism. | [(Slavotinek, Pua et al. 2017)](applewebdata://FA9EE35C-7824-4A91-9A1E-4D2B671FD335/#_ENREF_67) |
| THOC6 | 16 | *NO* | THO complex 6.This gene encodes a subunit of the multi-protein THO complex, which is involved in coordination between transcription and mRNA processing. The THO complex is a component of the TREX (transcription/export) complex, which is involved in transcription and export of mRNAs. | [(Accogli, Scala et al. 2018)](applewebdata://FA9EE35C-7824-4A91-9A1E-4D2B671FD335/#_ENREF_4) |
| ZBTB20 | 3 | *Yes* | Zinc finger and BTB domain containing 20.This gene, which was initially designated as dendritic cell-derived BTB/POZ zinc finger (DPZF), belongs to a family of transcription factors with an N-terminal BTB/POZ domain and a C-terminal DNA-bindng zinc finger domain. The BTB/POZ domain is a hydrophobic region of approximately 120 aa which mediates association with other BTB/POZ domain-containing proteins. This gene acts as a transcriptional repressor and plays a role in many processes including neurogenesis, glucose homeostasis, and postnatal growth. | [(Cleaver, Berg et al. 2019)](applewebdata://FA9EE35C-7824-4A91-9A1E-4D2B671FD335/#_ENREF_18) |
| ZMPSTE24 | 1 | NO | Zinc metallopeptidase STE24.This gene encodes a member of the peptidase M48A family. The encoded protein is a zinc metalloproteinase involved in the two step post-translational proteolytic cleavage of carboxy terminal residues of farnesylated prelamin A to form mature lamin A. | [(Shastry, Simha et al. 2010)](applewebdata://FA9EE35C-7824-4A91-9A1E-4D2B671FD335/#_ENREF_65) |

**Supplementary table 3. The identified patients with pathogenic variants in known genes. Note, only the genes that were involved in two cases were counted. The genome is based on hg38. "U" and "B" in anonymized sample IDs indicate "unilateral" and "bilateral" cryptorchidism, respectively. “GAF” and “EAF” show the “Global allele frequency in genomAD” and “East Asian allele frequency in genomAD”, respectively.**

| **ID** | **gene** | **Site** | **HGVS** | **Deleterious Impacts** | **GAF** | **EAF** |
| --- | --- | --- | --- | --- | --- | --- |
| U106 | BCORL1 | X-130028727-G-A | NC_000023.11:g.130028727G>A;NP_001171701.1:p.(G1391R) | SIFT,PolyPhen,LIST-S2,M-CAP,MutationTaster,fathmm-MKLcoding | 0.0001815 | 0.005439 |
| U37 | BCORL1 | X-130028727-G-A | NC_000023.11:g.130028727G>A;NP_001171701.1:p.(G1391R) | SIFT,PolyPhen,LIST-S2,M-CAP,MutationTaster,fathmm-MKLcoding | 0.0001815 | 0.005439 |
| U50 | BCORL1 | X-130028727-G-A | NC_000023.11:g.130028727G>A;NP_001171701.1:p.(G1391R) | SIFT,PolyPhen,LIST-S2,M-CAP,MutationTaster,fathmm-MKLcoding | 0.0001815 | 0.005439 |
| U9 | BCORL1 | X-130028727-G-A | NC_000023.11:g.130028727G>A;NP_001171701.1:p.(G1391R) | SIFT,PolyPhen,LIST-S2,M-CAP,MutationTaster,fathmm-MKLcoding | 0.0001815 | 0.005439 |
| U60 | KDM6A | X-45069958-A-C | NC_000023.11:g.45069958A>C;NP_001278345.1:p.(K775T) | ClinPred,LIST-S2,M-CAP,MutationTaster,fathmm-MKLcoding | NA | NA |
| U58 | KDM6A | X-45110152-A-G | NC_000023.11:g.45110152A>G;NP_001278344.1:p.(D1412G) | BayesDelnoAF,LIST-S2,LRT,M-CAP,MutationTaster,PROVEAN,fathmm-MKLcoding | NA | NA |
| U51 | UBA1 | X-47212835-A-T | NC_000023.11:g.47212835A>T;NP_003325.2:p.(Y873F) | SIFT,PolyPhen,ClinPred,LIST-S2,LRT,M-PolyPhen,CAP,MetaRNN,MutationTaster,PROVEAN,fathmm-MKLcoding | NA | NA |
| U60 | UBA1 | X-47214843-G-C | NC_000023.11:g.47214843G>C;NP_695012.1:p.(V1031L) | ClinPred,LIST-S2,M-CAP,MutationTaster,fathmm-MKLcoding | NA | NA |
| U49 | USP9Y | Y-12722148-G-A | NC_000024.10:g.12722148G>A;NP_004645.2:p.(E96K) | SIFT,PolyPhen,LIST-S2,M-CAP,MetaRNN,PROVEAN,fathmm-MKLcoding | NA | NA |
| U50 | USP9Y | Y-12722148-G-A | NC_000024.10:g.12722148G>A;NP_004645.2:p.(E96K) | SIFT,PolyPhen,LIST-S2,M-CAP,MetaRNN,PROVEAN,fathmm-MKLcoding | NA | NA |
| U109 | USP9Y | Y-12739592-C-G | NC_000024.10:g.12739592C>G;NP_004645.2:p.(S462C) | SIFT,PolyPhen,LIST-S2,M-CAP,PROVEAN,fathmm-MKLcoding | NA | NA |
| U71 | USP9Y | Y-12739592-C-G | NC_000024.10:g.12739592C>G;NP_004645.2:p.(S462C) | SIFT,PolyPhen,LIST-S2,M-CAP,PROVEAN,fathmm-MKLcoding | NA | NA |
| U73 | USP9Y | Y-12739592-C-G | NC_000024.10:g.12739592C>G;NP_004645.2:p.(S462C) | SIFT,PolyPhen,LIST-S2,M-CAP,PROVEAN,fathmm-MKLcoding | NA | NA |
| U13 | USP9Y | Y-12739592-C-G | NC_000024.10:g.12739592C>G;NP_004645.2:p.(S462C) | SIFT,PolyPhen,LIST-S2,M-CAP,PROVEAN,fathmm-MKLcoding | NA | NA |

Note: the other known genes without qualified candidate variants including: RXFP2, ABCD4, ABL1, ACADVL, ACTA2, ACTB, ACTG2, ACTL6A, ACTL6B, ADARB1, ADAT3, ADNP, AEBP1, AFF4, AGO2, AKR1C2, AKR1C4, ALDH18A1, ALG12, ALG8, ALKBH8, ALX4, AMH, AMHR2, ANAPC1, ANK1, ANKLE2, ANKRD11, ANKRD17, ANOS1, AP1S2, APC2, ARID1A, ARID1B, ARID2, ARL6, ARNT2, ARVCF, ARX, ASH1L, ASXL3, ATAD3A, ATN1, ATP6V0A2, ATP6V1A, ATP6V1E1, ATPAF2, ATR, ATRX, AUTS2, AXL, B3GALNT2, B3GLCT, B4GALT7, B4GAT1, B9D1, B9D2, BAP1, BAZ1B, BBIP1, BBS1, BBS10, BBS12, BBS2, BBS4, BBS5, BBS7, BBS9, BCAS3, BCL7B, BDNF, BICRA, BIN1, BLM, BMP4, BPTF, BRAF, BRCA1, BRCA2, BRD4, BRF1, BRIP1, BUB1B, BUD23, C2CD3, CASZ1, CBL, CC2D2A, CCBE1, CCDC141, CCDC174, CCDC22, CCDC32, CD96, CDC42, CDC45, CDC6, CDH11, CDH2, CDK8, CDKN1C, CDON, CDT1, CEP120, CEP152, CEP19, CEP290, CFAP418, CHD3, CHD4, CHD7, CHRM3, CHRNG, CHST14, CILK1, CITED2, CKAP2L, CLCN3, CLCN4, CLIP2, CLP1, CNOT3, COG1, COG5, COL3A1, COL4A1, COLEC10, COLEC11, COMT, CPE, CPLANE1, CPLX1, CPSF3, CREBBP, CRPPA, CSPP1, CTBP1, CTCF, CWC27, CYB5A, CYP11A1, CYP17A1, CYP19A1, CYP21A2, DACT1, DAG1, DAZ1, DAZ2, DAZ3, DAZ4, DCC, DDB2, DDX59, DDX6, DGCR2, DGCR6, DGCR8, DHCR7, DHDDS, DHODH, DHX37, DIS3L2, DKC1, DLG4, DLK1, DLL3, DLX4, DMRT3, DNAJC19, DNAJC21, DNAJC30, DNM2, DNMT3A, DOCK3, DOK7, DPAGT1, DPF2, DPYSL5, DSE, DUSP6, DVL1, DVL3, DYNC2H1, DYNC2I1, DYNC2I2, DYNC2LI1, DYRK1A, EBF3, EBP, EED, EFNB1, EHMT1, EIF2S3, EIF4H, EIF5A, ELN, EMC10, EMG1, EP300, ERCC1, ERCC2, ERCC3, ERCC4, ERCC5, ERCC6, ERCC8, ESCO2, ESS2, EVC, EVC2, EXT2, EZH2, FAM149B1, FANCA, FANCC, FANCD2, FANCE, FANCF, FANCG, FANCI, FANCL, FANCM, FARS2, FAT4, FBLN1, FBN1, FBXW11, FDFT1, FEZF1, FGD1, FGF10, FGF17, FGF8, FGFR1, FGFR2, FGFR3, FGFRL1, FIG4, FKBP6, FKRP, FKTN, FLI1, FLNB, FLRT3, FLT4, FMR1, FRAS1, FREM2, FTO, FUZ, FXR1, FZD2, G6PC3, GABRD, GALT, GATA1, GATA4, GATA5, GATA6, GDF1, GFM2, GJA5, GK, GLE1, GLI1, GLI2, GLI3, GMNN, GMPPB, GNB2, GNRH1, GNRHR, GP1BB, GPC4, GPC6, GPR161, GREAT, GRIA2, GRIA3, GRIA4, GRIP1, GSC, GTF2E2, GTF2H5, GTF2I, GTF2IRD1, GTF2IRD2, H19, H19-ICR, HBA1, HBA2, HDAC2, HDAC8, HERC2, HES7, HESX1, HIBCH, HIRA, HMGA2, HNF1B, HNRNPK, HOXD13, HPDL, HPSE2, HRAS, HS2ST1, HS6ST1, HSD17B3, HSD3B2, HSPA9, HSPG2, HTT, HUWE1, HYLS1, HYMAI, IER3IP1, IFT172, IFT27, IFT74, IFT80, IGBP1, IGF2, IL17RD, INPPL1, INSL3, IPW, IRF6, IRX5, JAG1, JAM3, JARID2, JMJD1C, KANSL1, KAT5, KAT6A, KAT6B, KAT8, KCNAB2, KCNMA1, KCNQ1, KCNQ1OT1, KDM1A, KDM3B, KDM4B, KDM5B, KDM5C, KDM6B, KDR, KIAA0753, KIF7, KISS1, KISS1R, KLHL40, KMT2D, KMT2E, KMT5B, KRAS, LARGE1, LETM1, LFNG, LHCGR, LHX1, LHX4, LIG4, LIMK1, LMBR1, LMBRD2, LMNB2, LMOD1, LMX1B, LONP1, LRIG2, LSM1, LSS, LUZP1, LZTFL1, LZTR1, MAD2L2, MADD, MAF, MAGEL2, MAMLD1, MAN2C1, MAP2K1, MAP2K2, MAP3K1, MAP3K7, MAPK1, MAPK8IP3, MAPRE2, MASP1, MBD5, MBTPS2, MC2R, MCM5, MCTP2, MECP2, MED12, MED13, MED13L, MED27, MEG3, MEGF8, MESP2, METTL27, MINPP1, MKKS, MKRN3, MKRN3-AS1, MKS1, MLXIPL, MMP23B, MPLKIP, MRAP, MRAS, MRPS28, MTM1, MTMR14, MTOR, MUSK, MYF6, MYH11, MYH3, MYL11, MYLK, MYMK, MYOD1, MYRF, MYT1L, NAA10, NALCN, NDN, NDNF, NDP, NEDD4L, NELFA, NF1, NFIB, NFIX, NIPBL, NKAP, NKX2-5, NKX2-6, NNT, NONO, NOTCH2, NOTCH3, NPAP1, NR0B1, NR2F2, NR5A1, NRAS, NSD1, NSD2, NSMF, NSUN2, NTNG1, NTNG2, NUP88, NXN, OCA2, OCRL, ODC1, OFD1, OGT, ORC1, ORC4, ORC6, OTUD5, OTUD6B, OTX2, P450c17, PACS1, PACS2, PALB2, PALS1, PAX6, PAX7, PBX1, PDE4D, PDE6D, PDPN, PEX1, PEX10, PEX11B, PEX12, PEX13, PEX14, PEX16, PEX19, PEX2, PEX26, PEX3, PEX5, PEX6, PHACTR1, PHF6, PHF8, PHGDH, PHIP, PIEZO2, PIGG, PIGN, PIGS, PIK3C2A, PLAG1, PLAGL1, PLVAP, PNPLA6, POLD1, POLE, POLR1B, POLR1C, POLR1D, POLR3A, POLR3K, POMGNT1, POMGNT2, POMK, POMT1, POMT2, POR, POU3F3, PPP1CB, PPP1R12A, PPP1R15B, PRDM13, PRDM16, PRKAR1A, PRKCZ, PRMT7, PROK2, PROKR2, PSMD12, PTCH1, PTCH2, PTDSS1, PTPN11, PTPN23, PTPRF, PUS7, PWAR1, PWRN1, PYCR1, RAB18, RAB23, RAB3GAP1, RAB3GAP2, RAC1, RAC3, RAD21, RAD51, RAD51C, RAF1, RAPSN, RARB, RASA2, RBM10, RBMY1A1, RECQL4, RERE, RFC2, RFT1, RFWD3, RIN2, RIPK4, RIPPLY2, RIT1, RLIM, RNF135, RNF2, RNU4ATAC, ROBO1, ROR2, RORA, RPGRIP1, RPGRIP1L, RRAS, RRAS2, RREB1, RSPO2, RTL1, RTTN, RXYLT1, RYR1, SALL1, SAMD9, SATB1, SATB2, SCAPER, SCYL2, SDCCAG8, SEC23A, SEC24C, SEMA3A, SETD1A, SETD2, SETD5, SGPL1, SHOC2, SIAH1, SIM1, SIN3A, SIX6, SKI, SLC12A2, SLC16A2, SLC18A3, SLC19A2, SLC25A24, SLC26A2, SLC35D1, SLX4, SMAD4, SMARCA2, SMARCA4, SMARCB1, SMARCC2, SMARCD1, SMARCE1, SMC1A, SMC3, SMCHD1, SMOC1, SMS, SNORD115-1, SNORD116-1, SNRPN, SOS1, SOS2, SOX10, SOX11, SOX2, SOX4, SOX9, SPECC1L, SPEN, SPRED2, SPRY4, SPTBN1, SRA1, SRCAP, SRD5A2, SRY, STAC3, STAG1, STAR, STRA6, STT3A, STT3B, STX1A, STXBP1, SUFU, SUZ12, SVBP, SYNCRIP, SYNE1, TAC3, TACR3, TAF6, TANC2, TARS1, TASP1, TBC1D20, TBCE, TBCK, TBL1XR1, TBL2, TBR1, TBX1, TBX22, TBX3, TCF12, TCF20, TCF4, TCF7L2, TCOF1, TCTN1, TCTN2, TCTN3, TFAP2A, TGDS, THOC6, TINF2, TMEM107, TMEM216, TMEM231, TMEM237, TMEM270, TMEM67, TMEM70, TMEM94, TNRC6B, TOE1, TOGARAM1, TOPORS, TP63, TPM2, TRAPPC4, TRIM32, TRIP12, TRIP4, TRMT1, TRRAP, TSPY1, TSPYL1, TTC5, TTC8, TUBB, TWIST1, TXNDC15, TXNRD2, UBE2A, UBE2T, UBE4B, UBR1, UBR7, UFD1, UQCC2, USP7, VAC14, VAMP7, VANGL1, VPS35L, VPS37D, VPS50, WASF1, WDFY3, WDPCP, WDR11, WDR35, WDR37, WNT3, WNT5A, WNT7A, WT1, WWOX, XPA, XPC, XRCC2, XRCC4, XYLT2, YY1, ZBTB20, ZEB2, ZFPM2, ZMIZ1, ZMPSTE24, ZMYM2, ZNF699, ZSWIM6, FANCB, NCF1, SEMA3E, CDCA7, THOC2, VPS13B, CUL4B, DDX3Y, FLNA, GPC3, KLHL15, LAS1L, MID1, NPHP1, PORCN, RNF113A, RPL10, SOX3, STS, OPHN1, AR, BRWD3, POLA1

**Supplementary table 4. The identified patients with pathogenic variants in novel genes. Note, only the genes that were involved in five cases were counted. The genome is based on hg38. "U" and "B" in sample IDs indicate "unilateral" and "bilateral" cryptorchidism, respectively. “GAF” and “EAF” show the “Global allele frequency in genomAD” and “East Asian allele frequency in genomAD”, respectively.**

| **ID** | **SYMBOL** | **HGVS** | **Deleterious prediction** | **GAF** | **EAF** |
| --- | --- | --- | --- | --- | --- |
| U27 | ARSH | NC_000023.11:g.3024049G>C;NP_001011719.1:p.(E310D) | PolyPhen, FATHMM, M-CAP, MetaLR, MetaSVM | NA | NA |
| U64 | ARSH | NC_000023.11:g.3024093G>C;NP_001011719.1:p.(G325A) | SIFT,PolyPhen,BayesDel_addAF_pred,BayesDel_noAF_pred,ClinPred_pred,DEOGEN2_pred,FATHMM_pred,LIST-S2,M-CAP_pred,MutationTaster_pred,MetaLR_pred,MetaRNN_pred,MetaSVM,MutationAssessor_pred | NA | NA |
| B122 | ARSH | NC_000023.11:g.3033050C>G;NP_001011719.1:p.(P452A) | SIFT,PolyPhen,BayesDel_noAF,DEOGEN2_pred,FATHMM_pred,LIST-S2,M-CAP_pred,MutationTaster_pred,MetaLR,MetaSVM | 0.0001168 | 0.003384 |
| U127 | ARSH | NC_000023.11:g.3033050C>G;NP_001011719.1:p.(P452A) | SIFT,PolyPhen,BayesDel_noAF,DEOGEN2_pred,FATHMM_pred,LIST-S2,M-CAP_pred,MutationTaster_pred,MetaLR,MetaSVM | 0.0001168 | 0.003384 |
| U28 | ARSH | NC_000023.11:g.3033050C>G;NP_001011719.1:p.(P452A) | SIFT,PolyPhen,BayesDel_noAF,DEOGEN2_pred,FATHMM_pred,LIST-S2,M-CAP_pred,MutationTaster_pred,MetaLR,MetaSVM | 0.0001168 | 0.003384 |
| U42 | DMD | NC_000023.11:g.31774145C>T; LRG_199p1:p.(E2453K) | PolyPhen,ClinPred,M-CAP_pred,MutationTaster | NA | NA |
| U55 | DMD | NC_000023.11:g.32342234G>A;NP_004000.1:p.(R1926C) | SIFT,PolyPhen,LIST-S2,M-CAP_pred,MutationTaster | NA | NA |
| U114 | DMD | NC_000023.11:g.32346044G>C;NP_000100.2:p.(Q1821E) | M-CAP | NA | NA |
| U58 | DMD | NC_000023.11:g.32346044G>C;NP_000100.2:p.(Q1821E) | M-CAP | NA | NA |
| B130 | DMD | NC_000023.11:g.32454661C>G;NP_004001.1:p.? | MutationTaster | NA | NA |
| U70 | DMD | NC_000023.11:g.32472199A>G;NP_000100.2:p.(Y964H) | PolyPhen,ClinPred,LIST-S2,M-CAP,MetaRNN | NA | NA |
| U111 | DMD | NC_000023.11:g.32573783C>T; NP_000100.2:p.(D548N) | SIFT,FATHMM_pred,LIST-S2,M-CAP_pred,MutationTaster_pred,MetaLR,MetaSVM | 0.0002763 | 0.007839 |
| U106 | MAGEA4 | NC_000023.11:g.151924064G>A;NP_001011550.1:p.(A134T) | SIFT,LRT | 0.0002307 | 0.005042 |
| U36 | MAGEA4 | NC_000023.11:g.151924064G>A;NP_001011550.1:p.(A134T) | SIFT,LRT | 0.0002307 | 0.005042 |
| U8 | MAGEA4 | NC_000023.11:g.151924064G>A;NP_001011550.1:p.(A134T) | SIFT,LRT | 0.0002307 | 0.005042 |
| U94 | MAGEA4 | NC_000023.11:g.151924064G>A;NP_001011550.1:p.(A134T) | SIFT,LRT | 0.0002307 | 0.005042 |
| U27 | MAGEA4 | NC_000023.11:g.151924194A>T;NP_001373127.1:p.(Y177F) | PolyPhen,ClinPred,LRT,MetaRNN | NA | NA |
| U98 | SHROOM2 | NC_000023.11:g.9786618C>A;NP_001640.1:p.(R25S) | SIFT,M-CAP | NA | NA |
| U33 | SHROOM2 | NC_000023.11:g.9894590G>A;NP_001640.1:p.(D228N) | PolyPhen,LIST-S2 | 0.000223 | 0.0006812 |
| U55 | SHROOM2 | NC_000023.11:g.9937394C>T;NP_001307593.1:p.(P118L) | SIFT,M-CAP | 0.00009806 | 0.0008489 |
| B79 | SHROOM2 | NC_000023.11:g.9937537G>T;NP_001307593.1:p.(A166S) | SIFT,PolyPhen,LIST-S2,M-CAP_pred,MutationTaster | NA | NA |
| B81 | SHROOM2 | NC_000023.11:g.9937537G>T;NP_001307593.1:p.(A166S) | SIFT,PolyPhen,LIST-S2,M-CAP_pred,MutationTaster | NA | NA |

**Supplementary table 5. The two heterozygous variants in RPTN. The genome is based on hg38.**

| **SampleID** | **impact** | **HGVSc** | **East Asian Allele frequency** | **Latino Allele frequency** | **Global Allele frequency** |
| --- | --- | --- | --- | --- | --- |
| U125 | stop_gained | NC_000001.11:g.152155265G>A;NP_001116437.1:p.(Q612*) | 0.003272 | 0.00006544 | 0.000118 |
| U16 | stop_gained | NC_000001.11:g.152155265G>A;NP_001116437.1:p.(Q612*) | 0.003272 | 0.00006544 | 0.000118 |
| U46 | stop_gained | NC_000001.11:g.152155265G>A;NP_001116437.1:p.(Q612*) | 0.003272 | 0.00006544 | 0.000118 |
| U103 | stop_gained | NC_000001.11:g.152155265G>A;NP_001116437.1:p.(Q612*) | 0.003272 | 0.00006544 | 0.000118 |
| U125 | frameshift_variant | NC_000001.11:g.152156949del;NP_001116437.1:p.(P51Qfs*8) | 0.003274 | 0.00006544 | 0.000118 |
| U16 | frameshift_variant | NC_000001.11:g.152156949del;NP_001116437.1:p.(P51Qfs*8) | 0.003274 | 0.00006544 | 0.000118 |
| U46 | frameshift_variant | NC_000001.11:g.152156949del;NP_001116437.1:p.(P51Qfs*8) | 0.003274 | 0.00006544 | 0.000118 |
| U103 | frameshift_variant | NC_000001.11:g.152156949del;NP_001116437.1:p.(P51Qfs*8) | 0.003274 | 0.00006544 | 0.000118 |
