## Supplementary note for "Novel mutation leading to splice donor loss in a conserved site of *DMD* causes cryptorchidism"

**Pathogenic variants identification in known and novel genes**

The general framework analyzing data flows this.

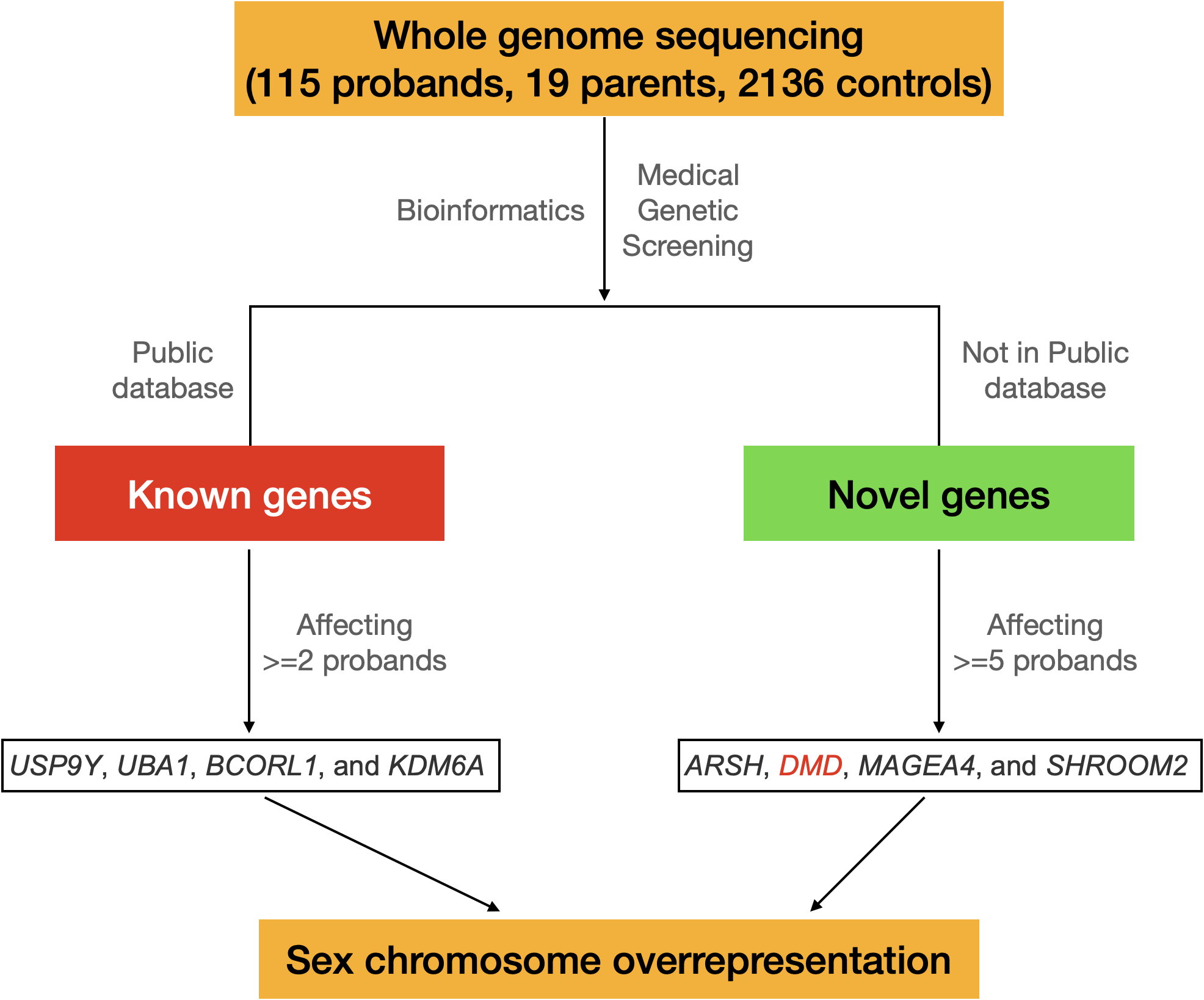

Scheme figure 1. The analyzing processes and major results for boys’ cryptorchidism based on whole genome sequencing data.

In detail, to determine rare pathogenic variants for both known and novel genes, we used a pipeline comprised of the co-segregation analysis, variant impact prediction, and common variants filtering by incorporating multiple software and databases, including SnpEff v5.1 ^1^, gnomAD v3.1.2 ^2^, 1000 genomes ^3^, PLINK v1.9 ^4^, VEP ^5^, SIFT ^6^, PolyPhen ^7^, LoFtools ^8^, BLOSUM ^9^, and dbNSFP ^10^. Only the variants with genetic consequence categories “HIGH” and “MODERATE” based on the VEP were included. The "HIGH" impact variants indicate disruptive effects on genes or chromosomes, such as the chromosome number variation, exon loss variant, frameshift variant, rare amino acid variant, splice acceptor variant, splice donor variant, start lost, stop gained, stop lost, and transcript ablation. The "MODERATE" impacts, such as inframe insertion, disruptive inframe insertion, inframe deletion, disruptive inframe deletion, missense variant, splice region variant, 3 prime UTR truncation, and 5 prime UTR truncation, may result in a non-disruptive but still primary protein sequence-altering effects.

Specifically, variant filtering and prioritization were performed with the following criteria for all variants per sample obtained from HaplotypeCaller in the GVCF mode: (a) The homozygous and heterozygous sites were adjusted with a threshold of reads proportions (RP). To increase the reliability of genotyping, only the alternative variants with RP scores over 85% were assigned as homozygous sites. The sites with RP scores between 15% and 85% were assigned as heterozygous. (b) To exclude sites with a high mutation rate, only biallelic sites were kept. (c) Only the variants with ten or more supporting reads were allowed. (d) For variants with "MODERATE" impacts, only those predicted to be deleterious or damaging by at least one algorithm implemented in the dbNSFP database were included. (e) Since the undescended testes naturally exist in certain species of Boreoeutheria and Afrotheria, for missense variants, we kept only the sites conserved at least in primate species. (f) Considering the phenotypic complexity of cryptorchidism, genes without expression in any reproductive system, at levels of tissue RNA, single-cell RNA, and protein, were screened using the HPA database ^11^, and removed. (g) Variants with allele frequencies higher than 0.01 registered in any population database were removed. (h) The candidate variants' quality was finally manually visualized with IGV to inspect the mapping quality, and variants within highly variable regions were removed. (i) The co-segregation between genotype and affected status was based on autosomal recessive, compound heterozygous, and X-linked recessive inheritance mode. (j) All variants in the final list were confirmed using the Sanger sequencing. (k) The significance of identified novel genes was further confirmed using multiple algorithms of burden tests in the RVTESTS package by incorporating variants from the “1000 genomes” database ^12^.

Following the annotation of candidate variants, the known and novel candidate genes can be recognized. The known genes are within a local “gene pool” related to cryptorchidism reported in PubMed literature (Supplementary Table 2) and the HPO database ^13^, which integrates three databases: Orphanet ^14^, DECIPHER ^15^, and OMIM ^16^. The cases with candidate rare pathogenic variants in known genes were filtered out during the identification of novel candidate genes. To make a rigorous identification, only the known genes affecting at least two cases and novel genes affecting at least five cases were included to estimate the diagnostic rate. To statistically test our newly identified candidate genes, we performed burden tests for rare variants with the RVTEST package ^12^, based on the methods of CMC test ^17^, CMC Fisher’s Exact test ^17^, CMC Wald test ^17^, Fp test ^18^, and Zeggini test ^19^.
